# Loss of symmetric cell division of apical neural progenitors drives *DENND5A*-related developmental and epileptic encephalopathy

**DOI:** 10.1101/2022.08.23.22278845

**Authors:** Emily Banks, Vincent Francis, Sheng-Jia Lin, Fares Kharfallah, Vladimir Fonov, Maxime Levesque, Chanshuai Han, Gopinath Kulasekaran, Marius Tuznik, Armin Bayati, Reem Al-Khater, Fowzan S. Alkuraya, Loukas Argyriou, Meisam Babaei, Melanie Bahlo, Behnoosh Bakhshoodeh, Eileen Barr, Lauren Bartik, Mahmoud Bassiony, Miriam Bertrand, Dominique Braun, Rebecca Buchert, Mauro Budetta, Maxime Cadieux-Dion, Daniel Calame, Heidi Cope, Donna Cushing, Stephanie Efthymiou, Marwa A. Elmaksoud, Huda G. El Said, Tawfiq Froukh, Harinder K. Gill, Joseph G. Gleeson, Laura Gogoll, Elaine S.-Y. Goh, Vykuntaraju K Gowda, Tobias B. Haack, Mais O. Hashem, Stefan Hauser, Trevor L. Hoffman, Jacob S. Hogue, Akimoto Hosokawa, Henry Houlden, Kevin Huang, Stephanie Huynh, Ehsan G. Karimiani, Silke Kaulfuß, G. Christoph Korenke, Amy Kritzer, Hane Lee, James R. Lupski, Elysa J. Marco, Kirsty McWalter, Arakel Minassian, Berge A. Minassian, David Murphy, Juanita Neira-Fresneda, Hope Northrup, Denis Nyaga, Barbara Oehl-Jaschkowitz, Matthew Osmond, Richard Person, Davut Pehlivan, Cassidy Petree, Lynette G. Sadleir, Carol Saunders, Ludger Schoels, Vandana Shashi, Rebecca C. Spillman, Varunvenkat M. Srinivasan, Paria N. Torbati, Tulay Tos, Undiagnosed Diseases Network, Maha S. Zaki, Dihong Zhou, Christiane Zweier, Jean-François Trempe, Thomas M. Durcan, Ziv Gan-Or, Massimo Avoli, Cesar Alves, Guarav K. Varshney, Reza Maroofian, David A. Rudko, Peter S. McPherson

## Abstract

Developmental and epileptic encephalopathies (DEEs) are a heterogenous group of epilepsies in which altered brain development leads to developmental delay and seizures, with the epileptic activity further negatively impacting neurodevelopment. Identifying the underlying cause of DEEs is essential for progress toward precision therapies. Here we describe a group of individuals with biallelic variants in *DENND5A* and determine that variant type is correlated with disease severity. We demonstrate that DENND5A interacts with MUPP1 and PALS1, components of the Crumbs apical polarity complex, which is required for both neural progenitor cell identity and the ability of these stem cells to divide symmetrically. Induced pluripotent stem cells lacking *DENND5A* fail to undergo symmetric cell division during neural induction and have an inherent propensity to differentiate into neurons, and transgenic *DENND5A* mice, with phenotypes like the human syndrome, have an increased number of neurons in the adult subventricular zone. Disruption of symmetric cell division following loss of *DENND5A* results from misalignment of the mitotic spindle in apical neural progenitors. A subset of DENND5A is localized to centrosomes, which define the spindle poles during mitosis. Cells lacking DENND5A orient away from the proliferative apical domain surrounding the ventricles, biasing daughter cells towards a more fate-committed state and ultimately shortening the period of neurogenesis. This study provides a mechanism behind *DENND5A*-related DEE that may be generalizable to other developmental conditions and provides variant-specific clinical information for physicians and families.

## Introduction

Developmental and epileptic encephalopathies (DEEs) are characterized by developmental delay and drug-resistant seizures. Abnormal neurodevelopmental outcomes are the result of both the underlying etiology and the negative impact of frequent epileptic activity on the developing brain^1^. Homozygous variants in *DENND5A* (DENN Domain Containing protein 5A), a gene encoding a protein expressed at high levels in the developing brain, have been linked to epileptic encephalopathy^2,3^, but the function of the protein during development is unknown. The conserved protein modules found in DENND5A and previously identified protein-protein interactions suggest its involvement in membrane trafficking and signal transduction.

DENND5A contains a DENN domain, an evolutionarily conserved protein module that functions as guanine nucleotide exchanges factors (activators) for Rab GTPases^4^, as well as additional protein modules known as RUN (RPIP8 [RaP2 interacting protein 8], UNC-14 and NESCA [new molecule containing SH3 at the carboxylterminus]) domains that also often interact with Rab family members^5^. Through the most N-terminal RUN domain (RUN1), DENND5A is an effector for the active form of Rab6^6–8^, and through the more C-terminal RUN domain (RUN2), DENND5A interacts with sorting nexin 1 (SNX1), a protein involved in protein trafficking between endosomes and the trans-Golgi network^9,10^. A role for DENND5A in cell division and polarity has been suggested in *in vitro* studies examining HeLa cells^11^ and a canine kidney model of cell polarity^12^, and *DENND5A* loss-of-function variants genetically predispose individuals to familial cutaneous melanoma due to SNX1-related protein trafficking deficits^13^. Whether and how cell division and polarity are affected during neurodevelopment with loss-of-function *DENND5A* variants has not been investigated.

The establishment and maintenance of apicobasal cell polarity is crucial in neural development. In early cortical development, apical cell membranes orient towards a central fluid-filled lumen^14^, which manifests as polarized neuroepithelial cells (NECs) or apical radial glia that respond to cerebrospinal fluid (CSF) dynamics via their apical membranes in the developing lateral ventricles^15,16^. Perturbations in genes encoding proteins that make up tight junctions (TJ) and adherens junctions (AJ), which define the apical plasma membrane and are often involved in cell division mechanisms, are associated with developmental defects consistent with DEE^17–20^. For example, pathogenic recessive variants in the gene encoding the TJ protein occuldin (*OCLN*) result in DEE due to defects in neural progenitor cell (NPC) symmetric cell division caused by impaired mitotic spindle formation and centrosome misalignment during mitosis^21,22^. Similarly, depletion of the TJ and AJ protein PALS1 (Protein Associated with Lin7 1) results in loss of symmetric cell division and enhanced cell cycle exit in NPCs^23^ and retinal neuroepithelia^24^ *De novo* variants in PALS1 have been linked to developmental delay and neurological abnormalities^19,23^.

Proteins comprising the apical membrane-defining Crumbs (Crb) complex in NECs include CRB2, MUPP1 (Multi-PDZ Protein 1, also known as MPDZ) or PATJ (Pals1-Associated Tight Junction Protein), and PALS1, and have been extensively studied for their roles in cell polarization and tissue development^25,26^. Most research on the Crb complex focuses on the version of the complex containing PATJ rather than MUPP1, and the only study directly comparing the function of these two highly similar proteins concludes that PATJ is essential but MUPP1 is dispensable for establishing and maintaining TJs^27^. *Ex vivo* studies, however, reveal significant deficits in ependymal^28,29^ and choroid plexus epithelial cells^30^ upon loss of MUPP1, including a complete loss of PALS1 expression in ependymal cells of *MUPP1* KO mice^29^. Although ependymal cells do not have TJs^31^ or stem cell properties^32^, they derive from TJ-containing neuroepithelial cells that gradually transition into radial glial cells that rely on abundant AJ protein expression to maintain their progenitor identity and ability to divide symmetrically^21,23,33,34^. Indeed, MUPP1 may differ from PATJ in that it preferentially stabilizes AJs over TJs^27^, which may suggest a radial glial-specific function for this complex during neural development.

Here we describe a cohort of 24 individuals from 22 families with biallelic *DENND5A* variants and determine their clinical presentations through phenotypic surveys answered by their treating clinicians, coupled with MRI analysis. We also demonstrate that DENND5A interacts with both PALS1 and MUPP1. To gain insight on the function of DENND5A, we employ mouse and zebrafish models of *DENND5A*-related DEE and perform *in-vitro* assays using *DENND5A* knockout (KO) induced pluripotent stem cells (iPSCs), and we show that disruption of symmetric cell division following loss of *DENND5A* results from misalignment of the mitotic spindle, pushing daughter cells toward a more fate-committed state and shortening the period of neurogenesis.

## Results

### Phenotypic characterization of individuals with biallelic *DENND5A* variants

Following our initial analysis of two families with homozygous variants in DENND5A^2^, we identified a cohort of 24 people (11 F, 13 M, mean age = 9.0 years, *SD* = 6.0) from 22 families with biallelic *DENND5A* variants. Thirty unique *DENND5A* variants were identified across the 14 homozygous and 10 compound heterozygous individuals. Seven members of the cohort have at least one additional variant flagged as potentially causative. Table 1 summarizes each person in the cohort, including their participant IDs, gene variant(s), predicted American College of Medical Genetics and Genomics (ACMG) variant interpretations, allele frequencies obtained from the gnomAD v2.1.1 dataset (https://gnomad.broadinstitute.org), seizure types and response to anti-seizure medications, occipitofrontal circumferences (OFCs), calculated scores corresponding to neurological and developmental phenotypes, and developmental outcomes. Pedigrees are available in Extended Data Figure 1 for participants 25-30, and some have affected family members not included in the cohort due to the unavailability of their clinical data. Pedigrees for participants 10, 15, and 16 were published previously^2^. None of the *DENND5A* point mutations in the cohort were found in the homozygous state among 140,000 individuals on gnomAD, a database that removes individuals affected by severe pediatric disease, indicating that biallelic pathogenic variants are likely incompatible with normal development. Twenty-five of the variants are found in the coding sequence, 2 are copy number variants (exon 1-14 duplication [NC_000011.9:(9171749_9172227)_(9316934_9321244)dup] and exon 1 deletion [NC_000011.10:g. 9262758_9268826del]), and 3 are intronic variants located in splice sites (splice donor variants c.2283+1G>T and c.949+1G>A, and polypyrimidine tract variant c.950-20_950-17delTTTT). The coding variants span the length of the protein including 9 in the DENN domain, 2 in the RUN1 domain, 6 in the PLAT domain, 4 in the RUN2 domain, and 4 in predicted linker regions between the folded modules (Fig. 1a).

**Figure 1:**
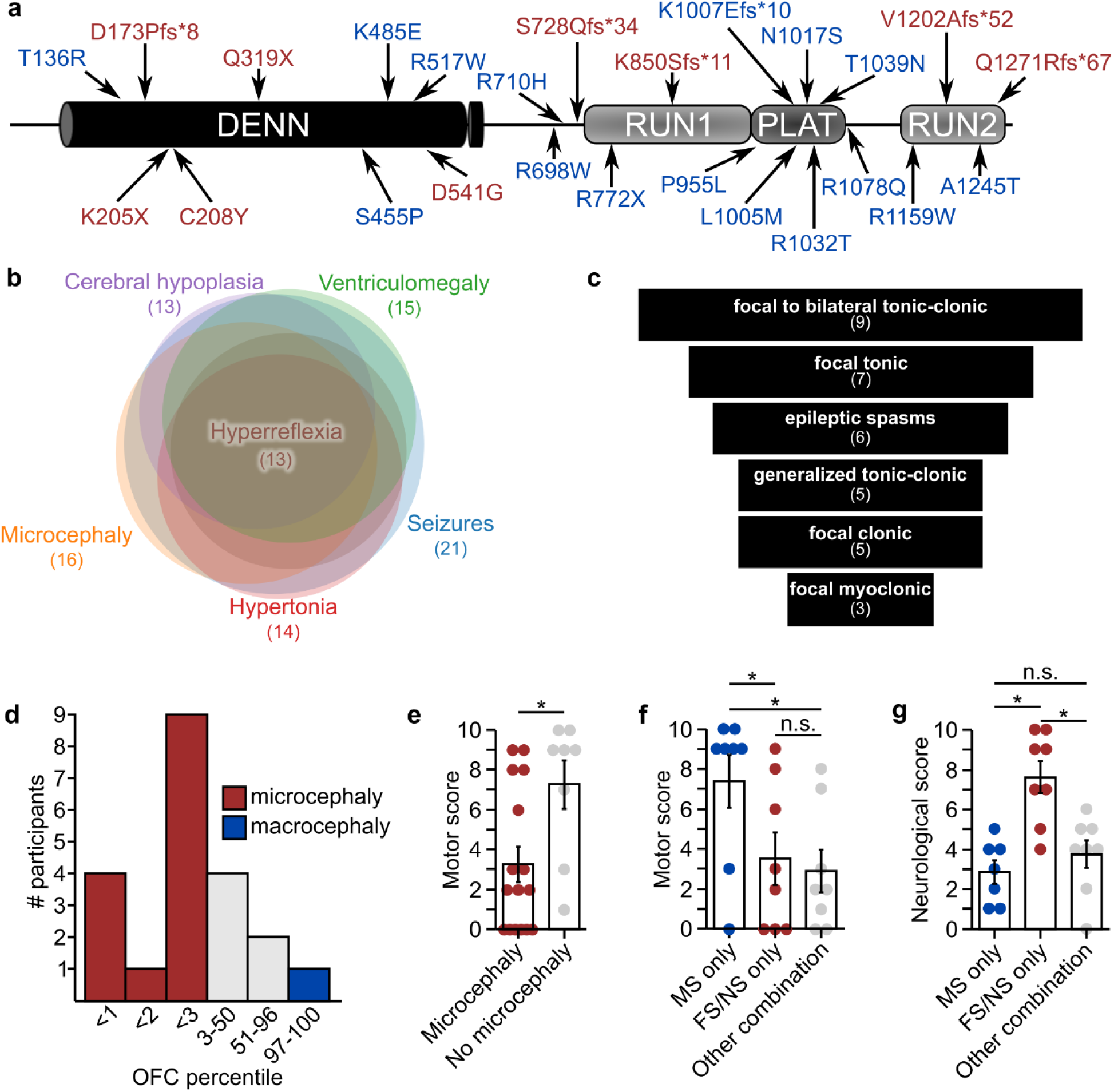
*DENND5A* loss of function variants influence neurodevelopment. **a,** Schematic of DENND5A protein with all coding sequence variants identified in the study. Red = found in homozygous individuals, blue = found in compound heterozygous individuals. **b,** Venn chart showing the number of people with biallelic *DENND5A* variants exhibiting the most frequently reported phenotypes and the degree of phenotypic overlap between cohort members. **c,** Funnel chart showing the most common seizure types present in the cohort. **d,** Histogram depicting the number of individuals in a given OFC percentile range. Note that the exact OFC percentile is not known in every case. **e,** Quantification of motor scores from *n* = 16 individuals with microcephaly and *n* = 8 individuals without microcephaly. Each dot represents one person. Data are mean ± SEM. **f,** Quantification of motor scores from *n* = 8 individuals with biallelic missense variants, *n* = 8 individuals with biallelic frameshift or nonsense variants, and *n* = 8 individuals with an allelic combination of frameshift, nonsense, missense, intronic, or copy number variants in DENND5A. Each dot represents one person. Data are mean ± SEM. **g,** Quantification of neurological scores from *n* = 8 individuals with biallelic missense variants, *n* = 8 individuals with biallelic frameshift or nonsense variants, and *n* = 8 individuals with an allelic combination of frameshift, nonsense, missense, intronic, or copy number variants in DENND5A. Each dot represents one person. Data are mean ± SEM.

**Table 1:**
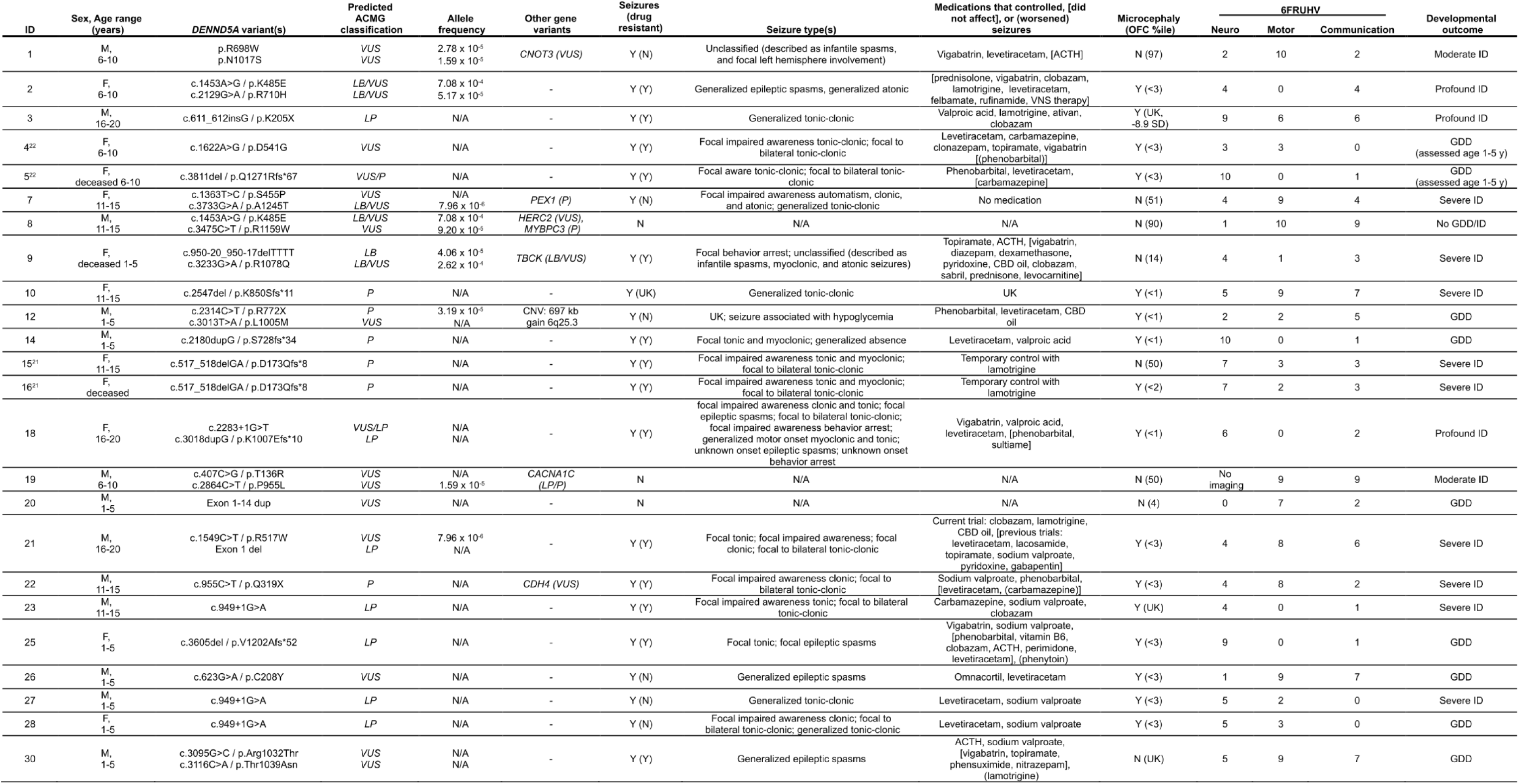
Clinical summary table indicating demographics, variant details with American College of Medical Genetics and Genomics classifications predicted by Franklin/Varsome, allele frequencies, and phenotype summaries for each individual in the study.

Complete phenotypic data for each member of the cohort is available in Source Data. The phenotypes observed in more than 50% of individuals with biallelic *DENND5A* variants were seizures (21/24), microcephaly (16/24), ventriculomegaly (15/24), hypertonia (14/24), cerebral hypoplasia (13/24), and hyperreflexia (13/24); Figure 1b). It is important to note that participants 8 (p.K485E/p.R1159W), 19 (p.P955L/p.T136R), and 20 (exon 1-14 dup) did not present with seizures. Participant 8 had a normal brain MRI and an autism spectrum disorder diagnosis requiring low levels of support, participant 20 exhibited global developmental delay with a normal brain MRI, and participant 19 presented with moderate intellectual disability but did not undergo neuroimaging. These observations suggest that these individuals do not have *DENND5A*-related DEE and that one or more of these variants may be benign or inherently less pathogenic.

Seizures were reported in 20/23 individuals, with an average age of onset of 4.8 months (*SD* = 5.9). Seizures typically onset within the first year of life with one patient experiencing their first seizure at 2 years of age. A funnel chart showing the frequencies of commonly reported seizure types is presented in Figure 1c. Focal to bilateral tonic-clonic are the most prominent seizure type, diagnosed in 9 individuals. Focal tonic seizures followed in 7 reported cases, 4 of whom were known to have impaired awareness. Among the 6 individuals presenting with epileptic spasms, 3 had a generalized onset, 2 had a focal onset, and 1 had an unknown onset. Generalized tonic-clonic seizures were reported in 5 individuals, as were focal clonic seizures. Three of the cases with focal clonic seizures had documented impaired awareness. Three individuals had focal myoclonic seizures, of which 2 cases had impaired awareness. In general, when the information was available, most focal seizures were accompanied by an impairment in awareness, with only one case retaining awareness. Seizures were generally drug resistant, but control was achieved in 6 cases with variable antiepileptic treatment. Anti-seizure medications that helped, did not affect, and worsened seizures can be seen for each case in Table 1.

All cases of microcephaly in the cohort were primary, with no cases of secondary microcephaly reported. Although microcephaly appears to be a major feature of *DENND5A*-related DEE, OFC percentiles ranged considerably (*M* = 18.5, *Mdn* = 2.9, *SD* = 30.2, *Min* = <1, *Max* = 97). A histogram depicting the distribution of known OFCs can be observed in Fig. 1d. One case (participant 1) of macrocephaly was reported, possibly secondary to their benign external hydrocephalus. Another case (participant 20) was noted to be “borderline” microcephalic with an OFC percentile of 4. Among the 7 individuals with normal OFCs, 6 underwent neuroimaging and 5 had clinically significant reductions in gray and/or white matter, indicating that neurodevelopment was compromised in most cases even when head circumference was within normal limits.

Failure to meet key developmental milestones was almost universal in the cohort, evidenced by the fact that all but one (participant 8; p.K485E/p.R1159W) presented with or had a history of global developmental delay. Among cohort members assessed after age 5, 9 had a severe intellectual disability (ID), profound ID was reported for 3 individuals, moderate ID was observed in 2 cases, and one participant (8) had no ID. Within the cohort, 15/24 (63%) were nonverbal, 7/24 (29%) were limited to single-word speech, and 2/24 (8%) could speak in sentences. Eye contact was present in 11 of 24 cases (46%). Eight of 24 (33%) could walk independently and 11/24 (46%) were able to reach for and grasp objects. Motor skills were assessed via a scoring system across the group (Supplementary Methods 1a), where a low score corresponds to no or minimal motor skills. Motor capabilities were more severely affected in those with microcephaly (*M*_Micro_ = 3.2, *M*_No micro_ = 7.2, *SD*_Micro_ = 3.5, *SD*_No micro_ = 3.4, two-tailed Mann-Whitney *U*, *Z* = −2.55, *p* = .011; Fig. 1e). Additionally, those with biallelic missense variants in *DENND5A* (*M* = 7.4, *SD* = 3.7) had significantly higher motor scores compared to those with either biallelic frameshift or nonsense variants (*M* = 3.5, *SD* = 3.7), as well as those with a combination of missense, frameshift, nonsense, intronic, or copy number variants (*M* = 2.9, *SD* = 3.0, Kruskal-Wallis *H* = 7.02, *p* = .03; Fig. 1f).

MRIs or computed tomography (CT) scans revealed abnormalities in 20 of the 23 cases that underwent imaging. Normal MRIs were reported for participants 8, 20 and 26. We devised a scoring system (Supplementary Methods 1b) to analyze the extent of neurological phenotypes across the group and found that variant type influences neurological phenotype severity, with more abnormalities in individuals with biallelic frameshift or nonsense variants (*M* = 7.6, *SD* = 1.6) compared to those with both biallelic missense variants (*M* = 2.8, *SD* = 1.6, *p* = .0004) or another combination of variant types (*M* = 3.8, *SD* = 1.9, *p* = .002, one-way ANOVA, *F*(2, 20) = [12.996], *p* = .0002; Fig. 1g). No significant difference in neurological score was observed between those with biallelic missense variants and those with a combination of missense, nonsense, frameshift, intronic, or copy number variants (*p* = .657).

Not all MR/CT images were made available, but all available images are presented in Figure 2 and Extended Data Figure 2. Raw MRI data from 5 cases and raw CT data from 1 case were analyzed by a pediatric neuroradiologist. Of these cases, two unrelated individuals (participants 5 and 14), both homozygous for *DENND5A* frameshift variants, showed a “complete” phenotype and had an interesting combination of neuroanatomical abnormalities. These include severe dysgenesis of the basal ganglia with an indistinct and dysplastic thalamic transition, diencephalic-mesencephalic junction dysplasia, and cortical malformations, particularly with pachygyria involving the occipital lobes, a reduced volume of the white matter with associated striatal and periventricular calcifications and ventriculomegaly, agenesis or severe dysplasia/hypoplasia of the corpus callosum, thin anterior commissure, and variable degrees of pontocerebellar hypoplasia (Fig. 2a-b). CT image analysis of another homozygous individual with severe DEE also revealed hypoplasia of the corpus callosum, mild cerebral hypoplasia, and lenticulostriate and periventricular calcifications (Extended Data Fig. 2a). MRIs analyzed from two compound heterozygous cases that exhibited severe DEE showed relatively mild neuroanatomical phenotypes (participants 2 and 18; Fig. 2c-d). Raw MRI data from additional compound heterozygous cases (participants 9 and 30) were not available, but isolated images revealed mild hypoplasia of the corpus callosum (Extended Data Fig. 2b) and ventriculomegaly (Extended Data Fig. 2c). Participant 8 with variants p.K485E/p.R1159W, who does not present with DEE, had a normal MRI with only mild inferior cerebellar vermis hypoplasia (Extended Data Fig. 2d), providing further evidence for the benign or less deleterious nature of p.R1159W, but not p.K485E, since the latter variant was found in an individual with severe DEE and mild neuroanatomical phenotypes (participant 2, p.K485E/p.R710H; Fig. 2c).

**Figure 2:**
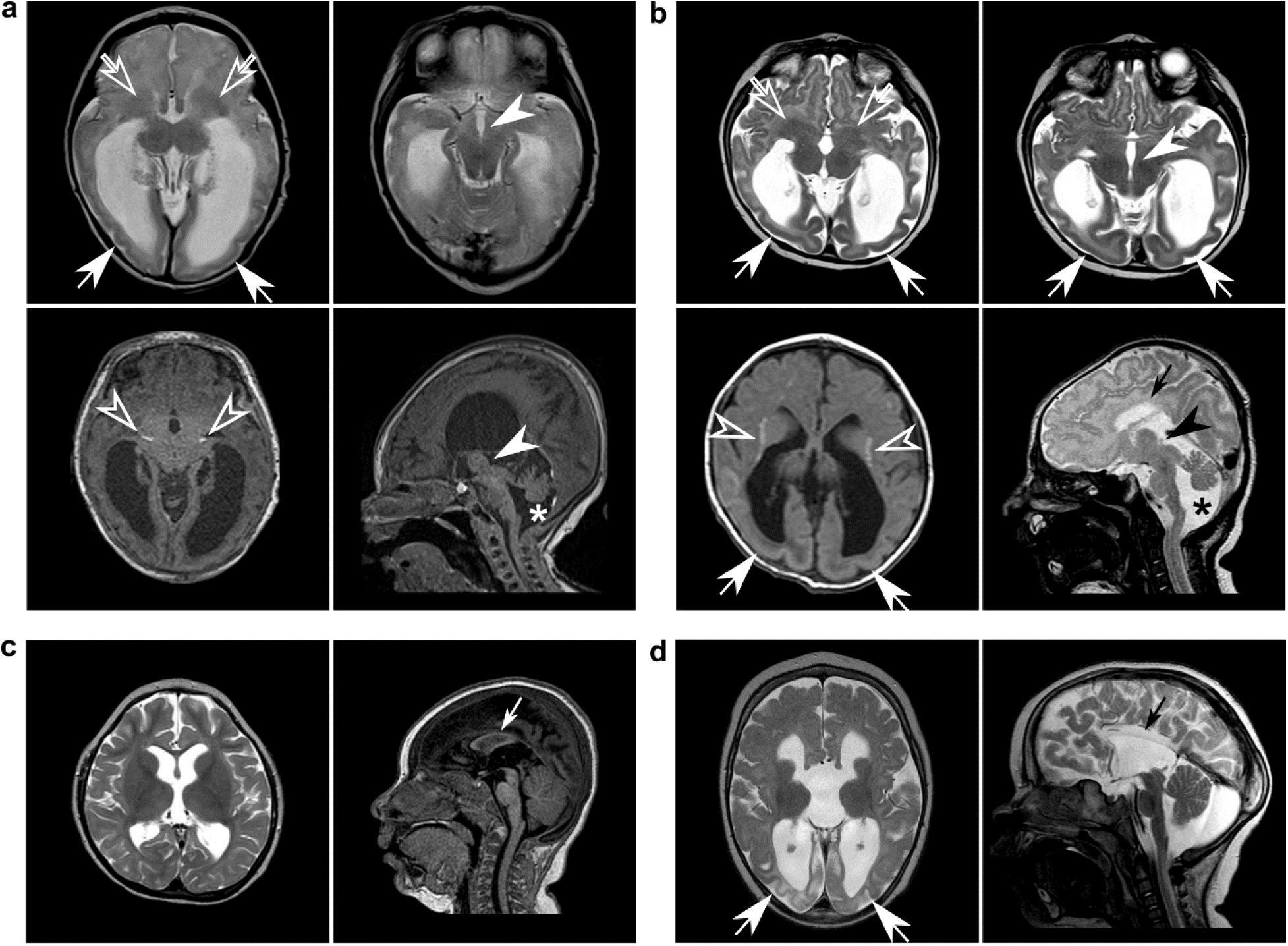
Cortical malformations, corpus callosum and anterior commissure dysgenesis, ventriculomegaly, basal ganglia dysgenesis, calcifications, and diencephalic/mesencephalic dysplasia are indicative of severe *DENND5A*-related DEE. Sample MRI slices from unrelated individuals with **a,** homozygous p.Q1271R*67 variants (participant 5); **b,** homozygous p.S728Qfs*34 variants (participant 14); **c,** compound heterozygous p.K485E/p.R710H variants (participant 2); and **d,** compound heterozygous c.2283+1G>T/p.K1007Efs*10 variants (participant 18) show many neuroanatomical phenotypes in common. Arrows = posterior gradient of pachygyria/lissencephaly; open arrows = severe basal ganglia dysmorphism; arrowheads = diencephalic/mesencephalic junction dysplasia; open arrowheads = periventricular, striatal, and diencephalic calcifications; small arrows = corpus callosum dysgenesis/agenesis; asterisks = cerebellar hypoplasia.

DENND5A protein levels were determined in cell lines derived from 5 cohort members: 3 from neural progenitor cells (NPCs) differentiated from iPSCs, and 2 from immortalized lymphoblasts, with homozygous and compound heterozygous as well as frameshift, nonsense, and missense variants represented. Positive controls (i.e. cells derived from healthy donors) were used in both experiments, and iPSC-derived cells were additionally compared against a negative control cell line in which KO of *DENND5A* was generated with CRISPR/Cas9 using guide RNAs targeting exon 4. NPCs express SOX1, SOX2 and Nestin, affirming their NPC identity (Extended Data Fig. 3). All patient-derived cells exhibit a reduction in DENND5A protein (Extended Data Fig. 4a-b), supporting that disease phenotypes are a result of protein loss of function; however, the KO-validated DENND5A antibody recognizes a region of the protein more C-terminal to the stop codon of p.K205X. RT-qPCR was thus performed, but a poor correlation between *DENND5A* mRNA and protein levels was observed as *DENND5A* mRNA expression from patient-derived cells did not differ significantly from controls (Extended Data Fig. 4c-d). This includes NPCs with homozygous p.K850Sfs*11 variants, where the antibody can determine that a ∼100 kDa truncated protein is not expressed because its epitope should still be detected, but mRNA levels were comparable to WT NPCs. When introduced into a FLAG-tagged DENND5A plasmid construct and overexpressed in HEK293T cells, p.K205X resulted in no detectable protein, indicating that even if a truncated protein is produced it is rapidly degraded (Extended Data Fig. 4e).

### Characterization of *DENND5A*-related DEE animal models

We established animal models in mice and zebrafish to study how biallelic pathogenic *DENND5A* variants affect development. A knock-in (KI) mouse model, homozygous for a frameshift variant (c.517_517delGA/p.D173Pfs*8) found in the proband of *DENND5A*-related DEE^2^ (participants 15 and 16) and conserved in mice (Extended Data Fig. 5a-b), exhibits anatomical and functional phenotypes consistent with those found in our human cohort. Immunoblotting indicated that full-length DENND5A protein is not expressed in KI mice (Fig. 3a); however, the validated DENND5A antibody has an epitope recognizing a region more C-terminal to the stop codon 8 amino acids past D173. RT-qPCR revealed a significant reduction in expression of *DENND5A* mRNA in KI mouse brains compared to WT (*M_WT_* = 1.0, *M_KI_* = 0.67, *SD_WT_* = 0.68, *SD_KI_* = 0.42, two-tailed Welch’s *t*(13.4) = 2.20, *p* = 0.046; Fig. 3b), suggesting nonsense-mediated RNA decay. Moreover, DENND5A protein containing the mutation and tagged at the N-terminus with FLAG is degraded when overexpressed in HEK-293T cells (Extended Data Fig. 4e), indicating that even if translated, the protein is likely degraded. *In vivo* 7T MRI scans revealed that *DENND5A* KI mice have significantly enlarged lateral ventricles (*M_WT_* = 4.8 mm^3^, *M_KI_* = 6.6 mm^3^, *SD_WT_* = 1.3, *SD_KI_* = 2.4, two-tailed Mann-Whitney U, *Z* = −2.117, *p* = .034), consistent with ventriculomegaly (Fig. 3c-d). KI mice also had lower mean and median relative brain sizes, but like our human cohort in which occipitofrontal circumference percentiles varied considerably, a high degree of variability was observed in the mice and the difference did not reach statistical significance (Fig. 3e). Finally, while spontaneous seizures were not observed in the KI mice, they show increased seizure susceptibility compared to WT when administered the potassium channel blocker 4-aminopyridine (*M_WT_* = 23.60, *M_KI_* = 11.67, *SD_WT_* = 2.97, *SD*_KI_ = 7.20, two-tailed *t*(9) = 3.445, *p* = 0.007; Fig 3f). A zebrafish model system was developed to study DENND5A during development. *dennd5a* mRNA was detected throughout embryonic development beginning from the first hour post-fertilization (hpf) (Extended Data Fig. 5c), and *in situ* hybridization revealed *dennd5a* expression at the two-cell stage (Fig. 3g). The mRNA was enriched in the central nervous system, retinal ganglion cells (RGCs), the sensory epithelium (otic vesicle, Ov), pharyngeal cephalic musculature (Cm), and the heart (H), as detected 24, 48, and 72 hpf (Fig. 3h-j). Biallelic mutations in *dennd5a* were introduced into zebrafish larvae using CRISPR/Cas9 (F_0_ KOs). Successful KO was confirmed by measuring *dennd5a* mRNA via RT-qPCR compared to Cas9 protein-injected controls (Extended Data Fig. 5d). F_0_ KOs show structural and behavioral perturbations consistent with *DENND5A*-related DEE including reduced head size and increased ventricle size (Fig. 3k-n). Moreover, the fish display behavioral phenotypes indicating neurological deficits, including altered locomotor activity during periods of light and dark and reductions in visual and acoustic startle responses (Extended Data Fig. 5e-h). Eye size is also reduced in F_0_ KOs (Extended Data Fig. 5i). Taken together, DENND5A appears to have conserved functions during development observable in multiple animal models.

**Figure 3:**
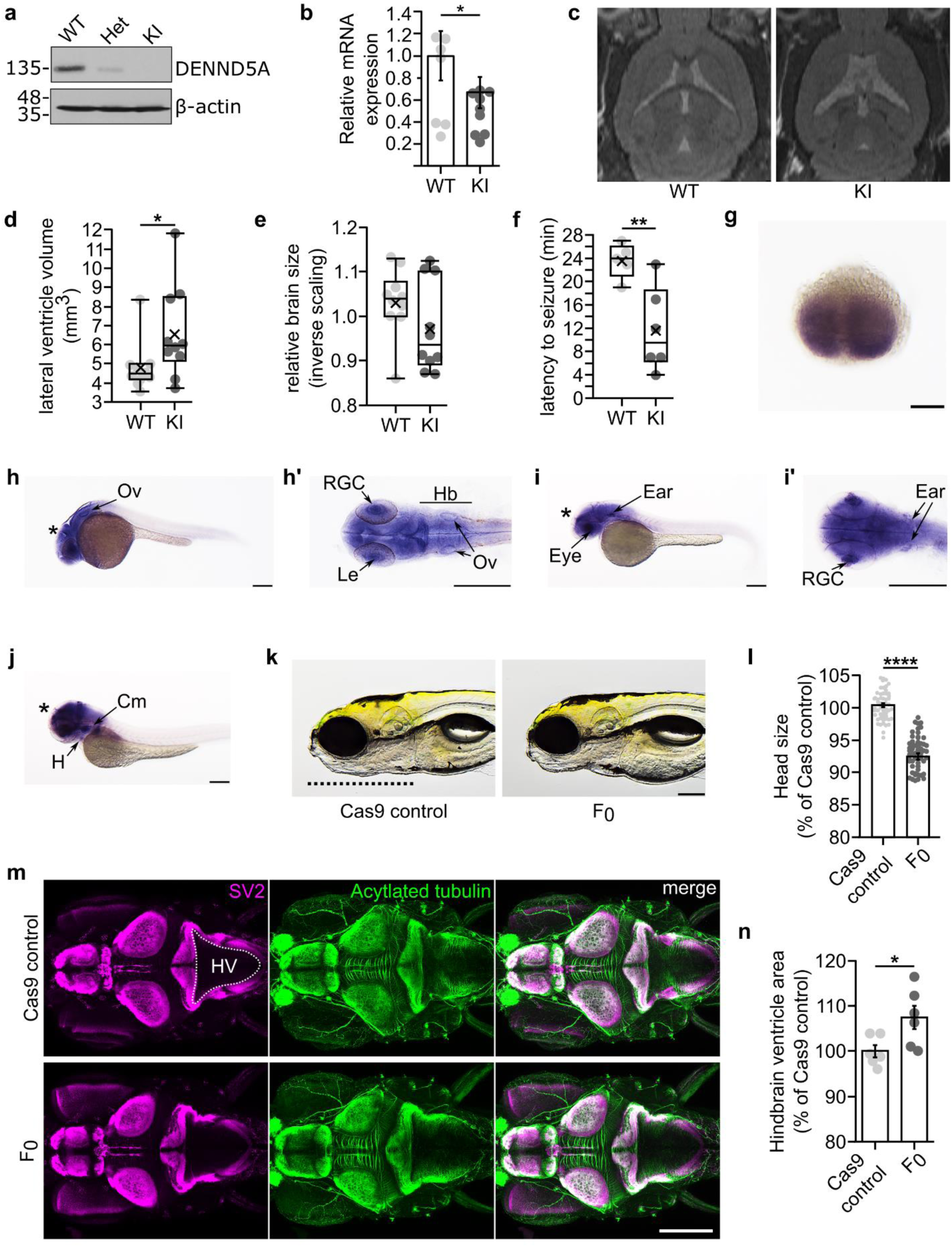
Animal models of *DENND5A*-DEE exhibit common phenotypes observed in the human cohort. **a,** Mice heterozygous (Het) for p.D173Pfs*8 express full-length DENND5A protein at half the levels compared to WT mice and homozygous knock-in (KI) mice express no full-length DENND5A protein. **b,** Relative brain *DENND5A* mRNA levels measured via RT-qPCR from *n* = 6 total mice. Experiments were performed in triplicate in 3 independent experiments. Error bars = SEM. **c,** Sample images of WT and KI *in vivo* 7T MRIs. **d,** Quantification of pooled lateral ventricle volumes obtained through segmenting *n* = 10 mouse MRIs. Each dot represents one animal. X = mean. **e,** Quantification of relative brain volumes measured using MRI data from *n* = 10 mice (*M_WT_* = 1.03, *Mdn_WT_* = 1.04, *M_KI_* = 0.97, *MdnKI* = 0.94, *SD_WT_* = 0.08, *SD_KI_* = 0.10, two-tailed Mann-Whitney U, *Z* = −1.361, *p* = .174). Each dot represents one animal. X = mean. **f,** Quantification of seizure latency after injection of 4-AP. Multiple independent experiments were performed with a total of *n* = 5 WT and *n* = 6 KI mice. Each dot represents one animal. X = mean. *(g-j)* Whole-mount *in situ* hybridization shows *dennd5a* mRNA expression at **g,** 0.75 hpf, **h,** 24 hpf, **i,** 48 hpf and **j,** 72 hpf. Asterisks = brain; Ov = otic vesicle; Le = lens; RGC = retinal ganglion cells; Hb = hindbrain; H = heart; Cm = cephalic musculature. Scale bar = 0.2 mm. **k,** Sample images of control and F_0_ KO zebrafish head size. Dotted line marks the length of the head used in quantification. Scale bar = 0.2 mm. **l,** Quantification of head size in *n* = 60 larvae analyzed via two-tailed Mann-Whitney U test (*Med*_Control_ = 100.432, *Med*_F0_ = 93.073, *SD*_Control_ = 2.316, *SD*_F0_ = 3.728; *Z* = −9.206, *p* < .0001). Each dot represents one larva. Data are mean ± SEM. **m,** Representative image of larva at 6 dpf immunostained with anti-SV2 (magenta) and anti-acetylated tubulin (green). Dorsal view, anterior to the left. HV = hindbrain ventricle. Dotted line outlines hindbrain ventricle area used in quantification. **N,** Quantification of hindbrain ventricle area in *n* = 6 larvae analyzed via two-tailed student’s *t*-test (*M*_Control_ = 100, *M*_F0_ = 107.502, *SD*_Control_ = 3.251, *SD*_F0_ = 6.386; *t*(10) = - 2.564, *p* = 0.028). Data are mean ± SEM. Each dot represents one larva.

### DENND5A interacts with protein components of apical polarity Crumbs complex

We screened for binding partners of DENND5A using affinity purification with multiple regions and domains of the protein as bait. Mass spectrometry of purified proteins revealed members of the Crb polarity complex, MUPP1 and PALS1, as major DENND5A binding partners, with the interaction mediated via a GST-tagged peptide flanking the missense variant R710H (Fig. 4a-b). This interaction was confirmed with proteins expressed in HEK293T cells (Fig. 4c).

**Figure 4:**
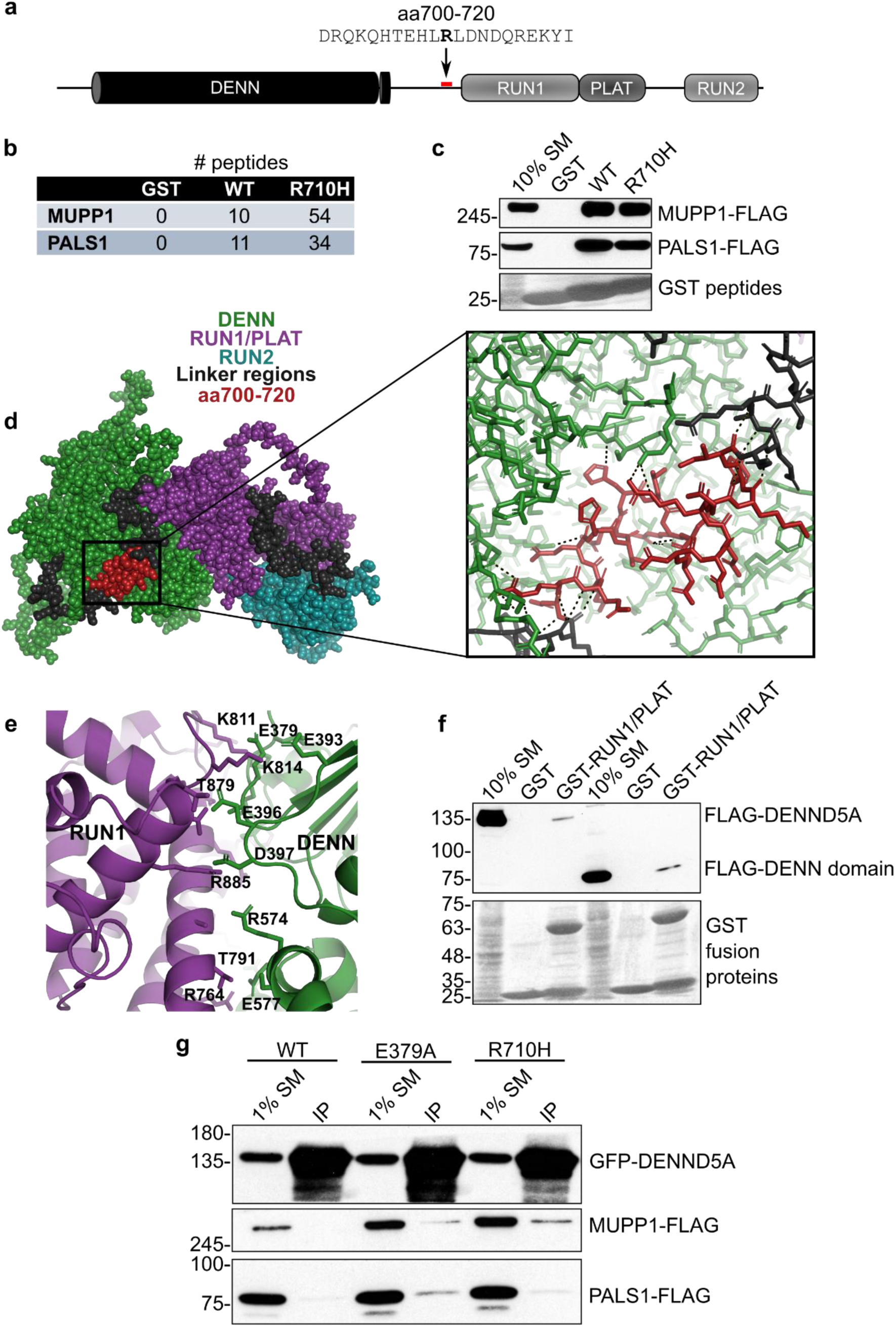
DENND5A interacts with polarity proteins MUPP1 and PALS1. **a,** A recombinant GST-tagged peptide containing amino acids 700-720 of human DENND5A sequence was generated for use in pulldown experiments. The bolded residue corresponds to Arg710 that is affected in the cohort (R710H). **b,** Table indicating the number of peptides corresponding to MUPP1 and PALS1 found bound to each GST fusion peptide used in the pulldown/mass spectrometry experiment. **c,** Overexpressed human MUPP1- and PALS1-FLAG bind to GST-tagged DENND5A peptides. **d,** Residues 700-720 are shown in red in a space-fill model (left) and magnified view (right) of the predicted DENND5A protein structure from AlphaFold. Dotted lines indicate hydrogen bonds. **e,** The interface between the DENN and RUN1 domains of DENND5A comprise many charged residues. **f,** GST pulldown experiments show that FLAG-DENN and GST-RUN1/PLAT physically interact. **g,** Co-immunoprecipitations between GFP-DENND5A and MUPP1- and PALS1-FLAG show that DENND5A only binds the polarity proteins when the intramolecular DENN-RUN1 interaction is disrupted.

We analyzed the predicted structure of DENND5A from AlphaFold^35,36^ and noted that residues R701, E707, H708, R710, and R716 involved in the interaction with the polarity proteins make hydrogen bonds with residues S10-A11, R273, R129, D598, and R716 of the DENN domain (Fig. 4d), suggesting a conformational change may be necessary to expose the binding site for MUPP1/PALS1. In fact, full-length DENND5A has limited interaction with these proteins (Fig. 4g). Because DENN domains are evolutionarily conserved protein modules that function as guanine nucleotide exchange factors for Rab GTPases^4^, we aligned the co-crystal structure of DENND1B and Rab35 with the predicted DENND5A structure and found that the predicted Rab enzymatic site in DENND5A is blocked by interactions between the DENN and RUN1/PLAT (Polycystin-1, Lipoxygenase, Alpha-Toxin) domains (PDB: 3TW8^37^; Extended Data Fig. 6a). In contrast, a conformational change is not necessary for the known DENND5A binding partner GTP-Rab6 to bind the RUN1 domain (PDB: 3CWZ^7^; Extended Data Fig. 6b), suggesting that two functional conformations are both possible and necessary.

We next examined the intramolecular interactions blocking MUPP1/PALS1 binding in the AlphaFold structure of DENND5A and observed a cluster of highly charged residues at the interface of the DENN and RUN1 domains (Fig. 4e), suggesting that the structure can be biochemically manipulated to expose both the Rab and Crb complex binding sites. To confirm this intramolecular interaction experimentally, we performed a pull-down assay with the GST-RUN1/PLAT domains as bait in HEK293T lysate expressing either full-length FLAG-tagged DENND5A or the isolated DENN domain (aa1-680). Both DENND5A constructs bind the RUN1/PLAT domain, with a slightly stronger interaction with the isolated DENN domain (Fig. 4f). Moreover, the DENN domain interaction was impeded with charge masking from higher salt concentrations (Extended Data Fig. 6c). We then performed mutagenesis experiments targeting residues involved in the intramolecular interaction. Mutating E379 resulted in a weak but present interaction between full-length DENND5A and Crb complex proteins, implying that DENND5A molecules were skewed toward an open conformation (Fig. 4g). R710H is predicted to result in the loss of a salt bridge to D598 of the DENN domain, potentially destabilizing the closed structure (Extended Data Fig. 6d). Like what was observed upon disrupting the DENN/RUN1 interaction, introducing the patient variant R710H also resulted in increased binding to MUPP1-FLAG and PALS1-FLAG (Fig. 4g). We conclude that DENND5A binds MUPP1 and PALS1 in a conformation-dependent manner, and that R710H increases the likelihood that DENND5A will adopt an open configuration.

### Loss of DENND5A drives premature neuronal differentiation

Microcephaly in *PALS1* conditional KO mice is due to neural progenitors prematurely exiting the cell cycle and undergoing asymmetric neurogenic cell divisions instead of symmetric proliferative divisions, resulting in the depletion of the progenitor pool and premature neuronal differentiation^23^. Our discovery of an interaction between DENND5A and PALS1 implies a degree of shared function. We thus tested if DENND5A also regulates cell division and differentiation phenotypes. We observed that *DENND5A* KO NPCs grow slower than WT (Fig. 5a). Interestingly, a significant difference was observed even after 24 hours (*M_WT_* = 710, *M_KO_* = 621, *SD_WT_* = 108.8, *SD_KO_* = 72.4, two-tailed *t*(18) = 2.168, *p* = .044), likely due to an increased number of apoptotic cells observed after plating KO cells. WT NPCs then rapidly increased in number whereas the number of KO NPCs remained relatively stable, producing a significant difference after 48 (*M_WT_* = 1008, *M_KO_* = 614, *SD_WT_* = 135.6, *SD_KO_* = 65.30, two-tailed Welch’s *t*(12.96) = 8.30, *p* < .0001) and 72 hours (*M_WT_* = 1685, *M_KO_* = 683, *SD_WT_* = 351.6, *SD_KO_* = 35.09, two-tailed Mann-Whitney U, *Z* = −3.78, *p* < .0001).

**Figure 5:**
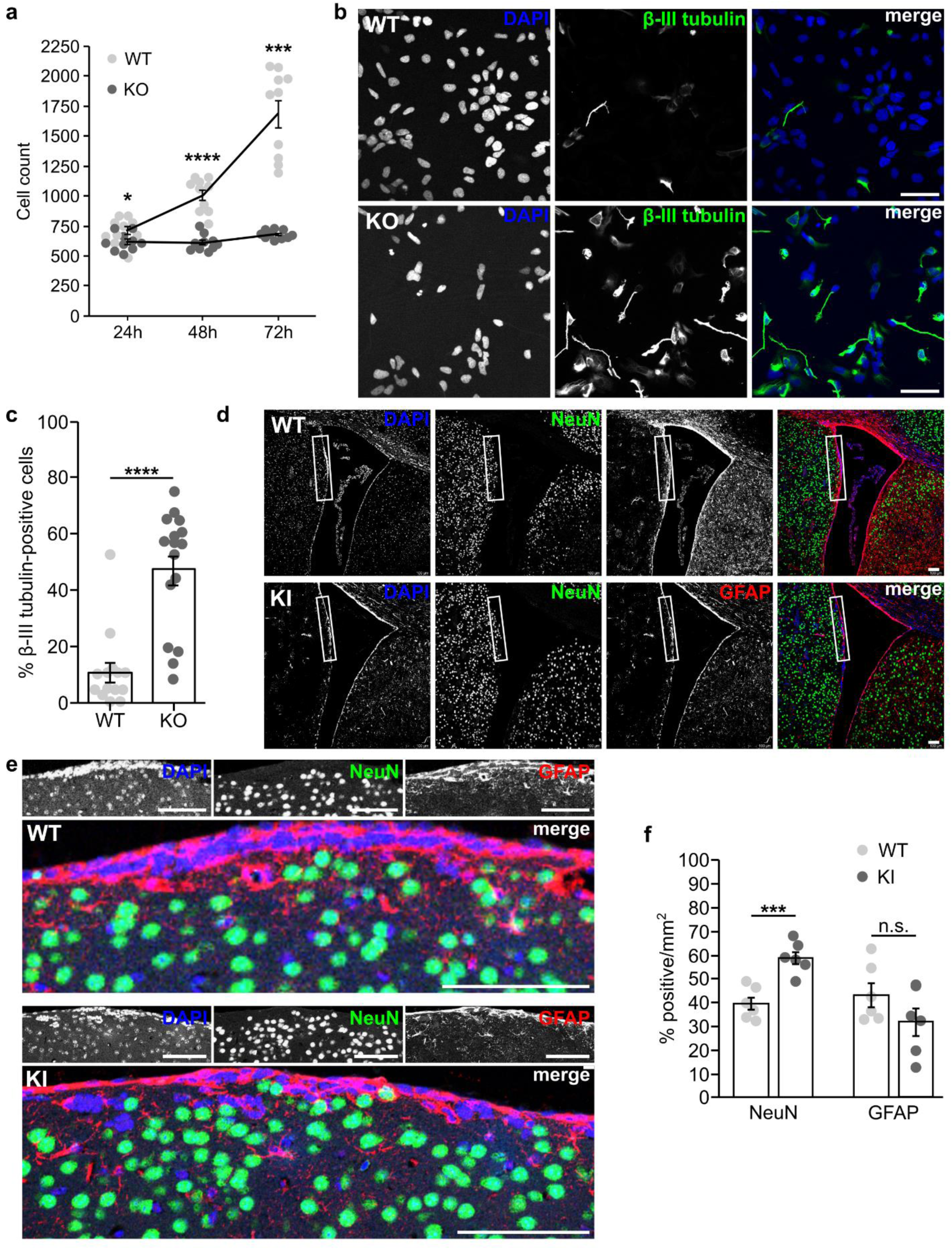
Loss of DENND5A results in premature neuronal differentiation. **a,** Graph showing the average number of NPCs counted per well of a 96-well plate 24, 48, and 72 hours after plating equal numbers of cells. Data are derived from 10 technical replicates from *n* = 2 independent experiments. Each dot represents the number of cells counted in one well. Error bars = SEM. **b,** Immunostaining of β-III tubulin (green) and DAPI (blue) in NPCs one day after plating into neural progenitor maintenance medium. Scale bar = 50 µm. **c,** Quantification of the percent of β-III tubulin-positive cells per field. A total of *n* = 2267 cells were analyzed from three independent experiments. Each dot represents the percentage calculated from one image. Data are means ± SEM. **d,** Immunostaining of GFAP (red), NeuN (green), and DAPI (blue) in the SVZ of adult mice. Scale bar = 100 µm. **e,** Close-up of the regions indicated in the insets in *(d)*. **f,** Quantification of the percentage of cells per mm^2^ labeled by NeuN or GFAP from a total of *n* = 4 mice. Each dot represents the percentage calculated from one image. Data are mean ± SEM.

Remarkably, after passaging newly-formed NPCs into neural progenitor maintenance medium, KO NPCs developed β-III tubulin-positive processes with neuronal morphology after one day, something rarely observed in WT NPCs (*M_WT_* = 10.67%, *M_KO_* = 47.29%, *SD_WT_* = 13.5, *SD_KO_* = 21.1, two-tailed Mann-Whitney U, *Z* = −3.991, *p* < .0001; Fig. 5b-c). To determine if this premature differentiation phenotype translates to complex organisms lacking *DENND5A*, we examined the adult mouse subventricular zone (SVZ), a region that normally retains GFAP-positive radial glia-like neural stem cells^38^ that are the primary source of newborn neurons in the adult SVZ^39^. KI mice have a significantly higher percentage of post-mitotic neurons expressing NeuN compared to WT (*M_WT_* = 39.6%, *M_KI_* = 58.8%, *SD_WT_* = 6.7, *SD_KI_* = 6.6, two-tailed *t*(10) = −4.981, *p* = 0.001; Fig. 5d-g). While there is also a reduction in the mean proportion of GFAP-positive cells in KI SVZs, it did not reach significance (*M_WT_* = 43.3%, *M_KI_* = 31.9%, *SD_WT_* = 12.3, *SD_KI_* = 14.1, two-tailed *t*(10) = 1.486, *p* = 0.168). Our results suggest that DENND5A expression promotes stemness and its loss permits cell cycle exit and premature differentiation.

### Loss of *DENND5A* misorients mitotic spindles

When the zebrafish homolog of *PALS1* is depleted from the developing retina, progenitor cells undergo asymmetric oblique cell divisions rather than symmetric horizontal divisions, detach from the apical ventricular surface, and differentiate prematurely^24^. We thus performed a neural rosette formation assay, a polarized *in vitro* model of early neural development, to assess the observed cell division and differentiation phenotypes to determine if the orientation of cell division is affected in *DENND5A* KO cells. WT and *DENND5A* KO iPSCs were plated at low density in neural induction medium and neural rosettes were allowed to form for up to 7 days. After 1 day *in vitro* (DIV), both WT and KO rosettes had maximal OCT4 expression, a marker of pluripotency, which rapidly declined by DIV 3 and was completely abolished by DIV 5 (Extended Data Fig. 7a). Expression of the NPC marker SOX2 was observed after 1 DIV, reached maximal levels at DIV 3, then its expression was slightly reduced and stabilized at DIV 5-7 (Extended Data Fig. 7b). This characterization is in line with rosettes generated from both iPSCs and embryonic stem cells using various neural induction protocols^34,40,41^. In general, there were many more WT rosettes formed per coverslip compared to KO. As in the NPC proliferation experiment, this may be due to the large amount of KO cell death observed at DIV 1, reducing the number of stem cells initially available for rosette formation. WT rosettes were considerably denser than KOs, but rosette diameter, lumen area, and lumen perimeter did not differ significantly (Extended Data Fig. 7c-e). PALS1 localized apically in both WT and KO rosettes (Extended Data Fig. 7f), suggesting DENND5A is not involved in trafficking MUPP1/PALS1 to the apical membrane. However, the axis of cell division in relation to the lumen differed (Fig. 6a). Because F-actin accumulates apically during rosette formation^42^ and outlines the cell borders of dividing cells, we used F-actin as a convenient marker of the apical surface. We measured the mitotic spindle angle, defined as the angle between the cleavage plane and the nearest apical surface, considering only cells with normally condensed chromatin and both centrosomes marked by γ-tubulin visible in the same plane. Although this exclusion criteria omitted many cells in WT rosettes dividing symmetrically along the z-plane or above the lumen (Extended Data Fig. 7g) as well as numerous observations of dividing cells with abnormally condensed chromatin in KO rosettes (Extended Data Fig. 7h), the spindle angle among cells dividing within WT (*M* = 57.1°, *Mdn* = 65.4°, *SD* = 25.9) and KO (*M* = 26.0°, *Mdn* = 20.1°, *SD* = 19.0) rosettes differed significantly according to a two-tailed Mann-Whitney U test (*Z* = −7.122*, p* < .0001; Fig. 6b). An overwhelming majority of KO cells divided with spindle angles <45° (Fig. 6c), indicating that *DENND5A* KO results in increased levels of oblique asymmetric cell divisions and the ability for apical progenitors to self-renew is severely compromised.

**Figure 6:**
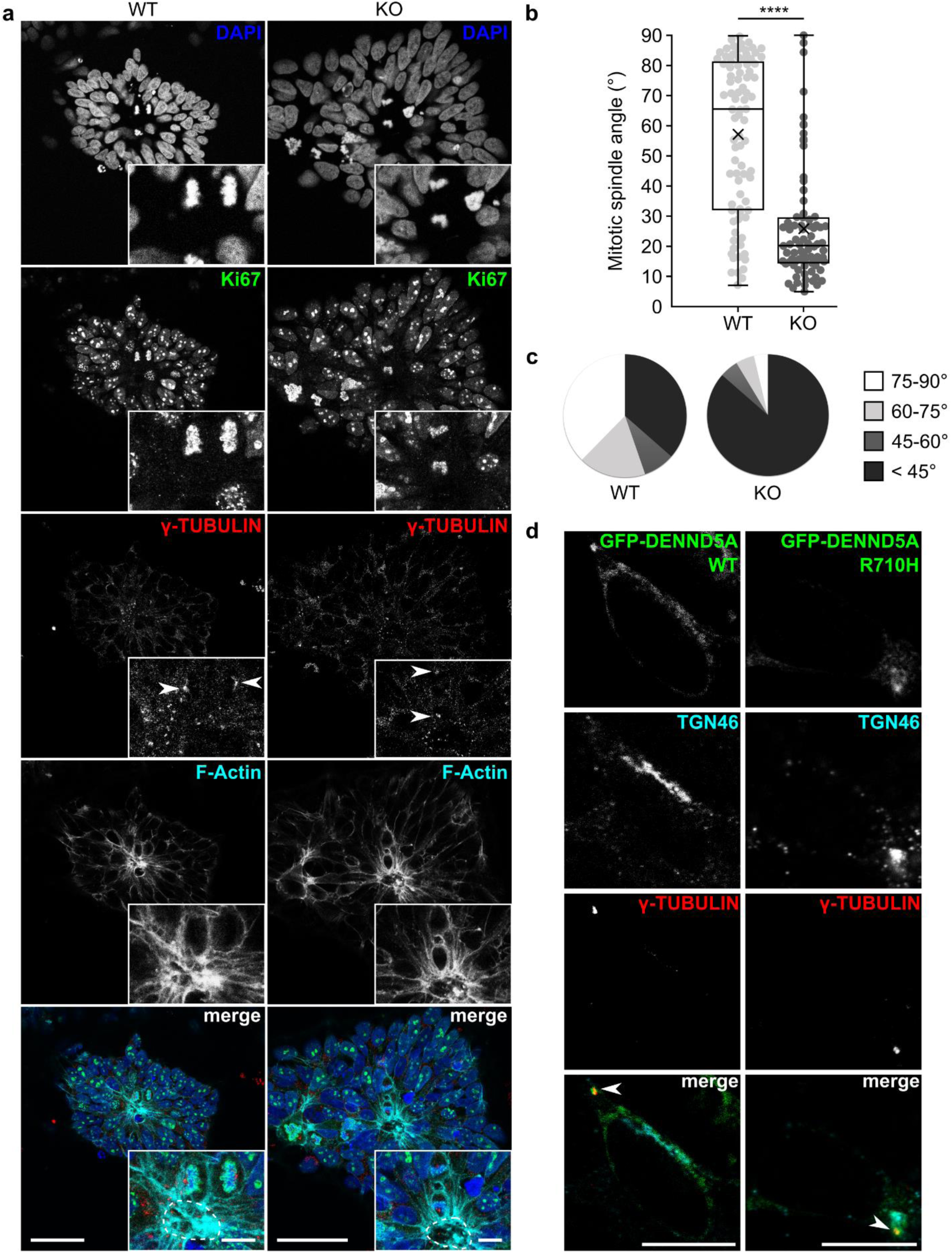
A neural rosette formation assay reveals abnormal mitotic spindle orientations upon loss of *DENND5A*. **a,** Sample images showing the orientation of apical progenitor cell division in WT and *DENND5A* KO rosettes. Green = Ki67, red = γ-tubulin, cyan = F-actin, blue = DAPI. Scale bars = 50 µm, inset = 10 µm. Dotted lines outline the F-actin positive lumen. **b,** Quantification of mitotic spindle angles measured from *n* = 85 WT and *n* = 81 KO dividing cells from 2 independent experiments. X = mean. **c,** Pie charts showing the proportion of dividing cells with mitotic spindle angles falling within various ranges. d, Overexpression of DENND5A in NPCs. Green = GFP-DENND5A, cyan = TGN46, red = γ-tubulin. Scale bars = 10 µm.

### A pool of DENND5A localizes to the centrosome

Alterations in cell division can result from changes in centrosome alignment, due to their nucleation of the astral microtubules of the mitotic spindle, which connect with the cell cortex and are the primary determining factor of daughter cell positioning^43^. We hypothesized that because almost all dividing cells with normally condensed chromatin in *DENND5A* KO rosettes divided with a perpendicular orientation, centrosome dynamics promoting parallel, symmetric divisions were compromised. To gain insight into the subcellular function of DENND5A, we examined the localization of GFP-DENND5A WT or R710H in NPCs. In addition to a Golgi localization, which is consistent with previous studies^11^, we identified that DENND5A also localized to γ-tubulin-positive centrosomes (Fig. 6d). No difference in localization was observed between WT and R710H constructs.

## Discussion

Here we present a cohort of individuals with biallelic variants in *DENND5A* leading to a new form of DEE. Some key similarities and differences between *DENND5A*-DEE and other monogenic DEEs described in the literature can be noted. Although none of the neuroanatomical features that we report in *DENND5A*-DEE are unique to DEEs, we are unaware of any other monogenic DEE in the literature with the same specific combination of features as those observed here. A study examining fetuses with diencephalic-mesencephalic junction dysplasias accompanied by developmental ventriculomegaly co-presented with corpus callosum dysgenesis and pontocerebellar hypoplasia, but no microcephaly was reported and their ventriculomegaly was associated with hydrocephalus due to aqueductal stenosis, a feature not identified in our cohort^44^. There is also a considerable degree of phenotypic overlap between our cohort and one comprised of patients with pathogenic *PCDH12* variants, but these patients lacked pachygyria and often co-presented with ophthalmic abnormalities, in contrast to what was observed here^45–47^. Gyral simplification and calcifications are observed in cases with recessive variants in the tight junction protein-encoding *OCLN* gene, but severe basal ganglia or diencephalic and mesencephalic dysplasias were not reported and polymicrogyria, a key feature of *OCLN* mutations, is not observed in our study^17^. Variants in tubulin genes also result in overlapping phenotypes^48^, but our cases lack the classic dysgyria pattern and instead are typically observed in the context of occipital pachygyria and differ in that our cases are associated with calcifications. Moreover, it seems that our cases have more severe basal ganglia abnormalities when compared with the typical imaging presentations associated with *TUBA1A*, *TUBB2A*, *TUBB2B*, *TUBB3*, and *TUBG1*^49–51^. Variants in *LIS1*, encoding a protein involved in neuronal migration, results most frequently in a posterior gradient of lissencephaly, without calcifications, and variable degrees of corpus callosum size including thin, normal, and thicker than normal tracts^52,53^. In contrast, corpus callosum volumes were either normal, reduced, or absent with no cases of increased volume in our cohort. Finally, there is a selective involvement of the cortex added to the presence of periventricular calcifications, a unique feature that brings cytomegalovirus-induced brain malformations into the differential but aligns with pseudo-TORCH syndrome diagnostic criteria in the absence of congenital infection^54,55^. These differences in neuroradiological features suggest an interesting genotype-phenotype relationship that warrants further study. Due to a small cohort size, limited neuroimaging data, and reliance on brief clinical reports for most cases, we were unable analyze all cases with the same degree of detail to determine the uniqueness of the genotype-phenotype relationship. Nonetheless, our radiological findings combined with clinical information in Table 1 and Source Data provide physicians with valuable information to communicate with families and treatment teams on a case-by-case basis.

For families in which one parent is aware they are a carrier of a pathogenic *DENND5A* variant, genetic counselors can recommend the other parent undergo genetic testing. When both parents are carriers, *in vitro* fertilization with preimplantation genetic diagnosis can be offered. However, the presence of *DENND5A* variants of unknown clinical significance does not necessarily equate to a devastating prognosis. 12% of our small cohort do not meet criteria for DEE or even experience seizures; p.R1159W, p.P955L, p.T136R, and/or exon 1-14 duplication may be benign variants or variants that have less impact on development. Additionally, we believe ACMG classifications for several variants should be updated to pathogenic or likely pathogenic, especially p.R517W, p.K485E, p.D541G and p.R710H, as these variants are found in individuals with brain abnormalities, severe intellectual disability, infantile seizure onset, and no other flagged gene variants. The identification of additional *DENND5A*-related DEE cases will prove valuable for future clinical and biological studies and thus improved treatment options and prognostic information for health care providers and families.

The identification of DENND5A as a polarity-related protein is consistent with previous clinical observations. Human cases with pathogenic variants in *CRB2* and *MUPP1* have been identified and exhibit ventriculomegaly and corpus callosum dysgenesis^18,28,56–58^, and patients with *PALS1* variants show global developmental delay, microcephaly, and sometimes seizures^19^. Moreover, the phenotypes observed in our *DENND5A*-related DEE mouse model overlap with those found in other mouse models targeting these proteins. *MUPP1* KO mice have enlarged lateral ventricles^30^ and the cortex in *PALS1* conditional KO mice, where PALS1 was selectively depleted from cortical progenitors, fails to develop, leading to microcephaly^23^. The weak strength of the interaction between open DENND5A and MUPP1/PALS1 may indicate that this occurs transiently in cells or only under specific circumstances. Indeed, the conformation-dependent nature of the interaction reflects the importance of these proteins remaining separate from each other under steady state conditions, and may reflect a molecular mechanism to regulate the balance of symmetric versus asymmetric cell division during mitosis. The biologically relevant mechanism for opening the DENND5A structure remains elusive, but possible candidates include posttranslational modifications or other currently unidentified protein-protein interactions. Future work should emphasize the mechanisms regulating the DENND5A conformational change and the role of its MUPP1/PALS1 interaction during cell division.

We have tested all commercial antibodies against DENND5A using several *DENND5A* KO cell lines and did not observe a specific signal via immunofluorescence, so the localization of endogenous DENND5A during cell division of apical progenitors is unknown. However, our overexpression studies indicate that a pool of DENND5A localizes to the centrosome during NPCs in interphase. No transfected cells in any other phase of the cell cycle were identified. One can speculate that DENND5A is involved in properly positioning centrosomes to align them parallel to the apical membrane during mitosis, ensuring that both daughter cells inherit apical determinants and remain in contact with the stem and progenitor cell biochemical niche found in the developing ventricle. This is indeed the case with the TJ and DEE-associated protein OCLN that binds directly to NuMa^21^, which under normal circumstances coordinates with dynein and plasma membrane-associated proteins to tightly align the mitotic spindle through pulling forces and tethering astral microtubules to the cell cortex^59–64^. DENND5A may also play a role in core mitotic spindle assembly, as abnormal chromatin condensation was observed in many dividing *DENND5A* KO NPCs. However, the loss of parallel spindle orientation relative to the apical surface in virtually all remaining cells suggests a major function in astral microtubule tethering. Mora-Bermúdez et al. showed that a decrease in the number of molecules that link astral microtubules to the apical cell cortex leads to a decrease in the number of apicobasal-specific astral microtubules, resulting in a weakened anchor between the spindle poles and the apical cell cortex and promoting an oblique or perpendicular spindle orientation^65^. We therefore hypothesize that DENND5A functions in this capacity; that the centrosomal pool of DENND5A radiates outward as apicobasal astral microtubules nucleate to link them with MUPP1/PALS1 at the apical cell cortex, promoting a planar spindle orientation. Future studies could quantify the number of apicobasal astral microtubules in WT versus *DENND5A* KO dividing cells within rosettes to investigate this hypothesis.

Alternatively, or perhaps concurrently, DENND5A may regulate the inheritance of MUPP1 and PALS1 to maintain a neural stem cell identity. DENND5A is also a Golgi-localized protein (Fig. 6d and ^11^) and the Golgi is confined to the apical process of apical progenitors in punctate stacks rather than a ribbon structure^65,66^. Golgi fragments containing DENND5A bound to PALS1/MUPP1 may thus ensure equal inheritance of apical determinants in both daughter cells of a dividing apical progenitor, affecting the resulting daughter cell fates.

Our study provides evidence for the involvement of *DENND5A* in two well-known processes implicated in primary microcephaly: centrosome positioning during cell division and premature NPC cell cycle exit and differentiation^67^. We propose a disease model, presented in Figure 7, in which *DENND5A*-related DEE is driven by a significant reduction in symmetric cell divisions during early development due to the misorientation of cells away from the proliferative apical domain of the ventricular zone. This results in an imbalance of signaling molecules from the stem and progenitor cell niche to each daughter cell and unequal inheritance of apical determinants such as MUPP1 and PALS1, biasing one daughter cell toward a more fate-committed state^68^. Ultimately, the period of neurogenesis is shortened which leads to microcephaly and/or observable abnormalities in gray and white matter structures. The reduced volume of neurons likely leads to compensatory ventriculomegaly, and improperly positioned prematurely-born neurons that do not undergo apoptosis may form aberrant synaptic contacts resulting in seizures that can further adversely affect development.

**Figure 7:**
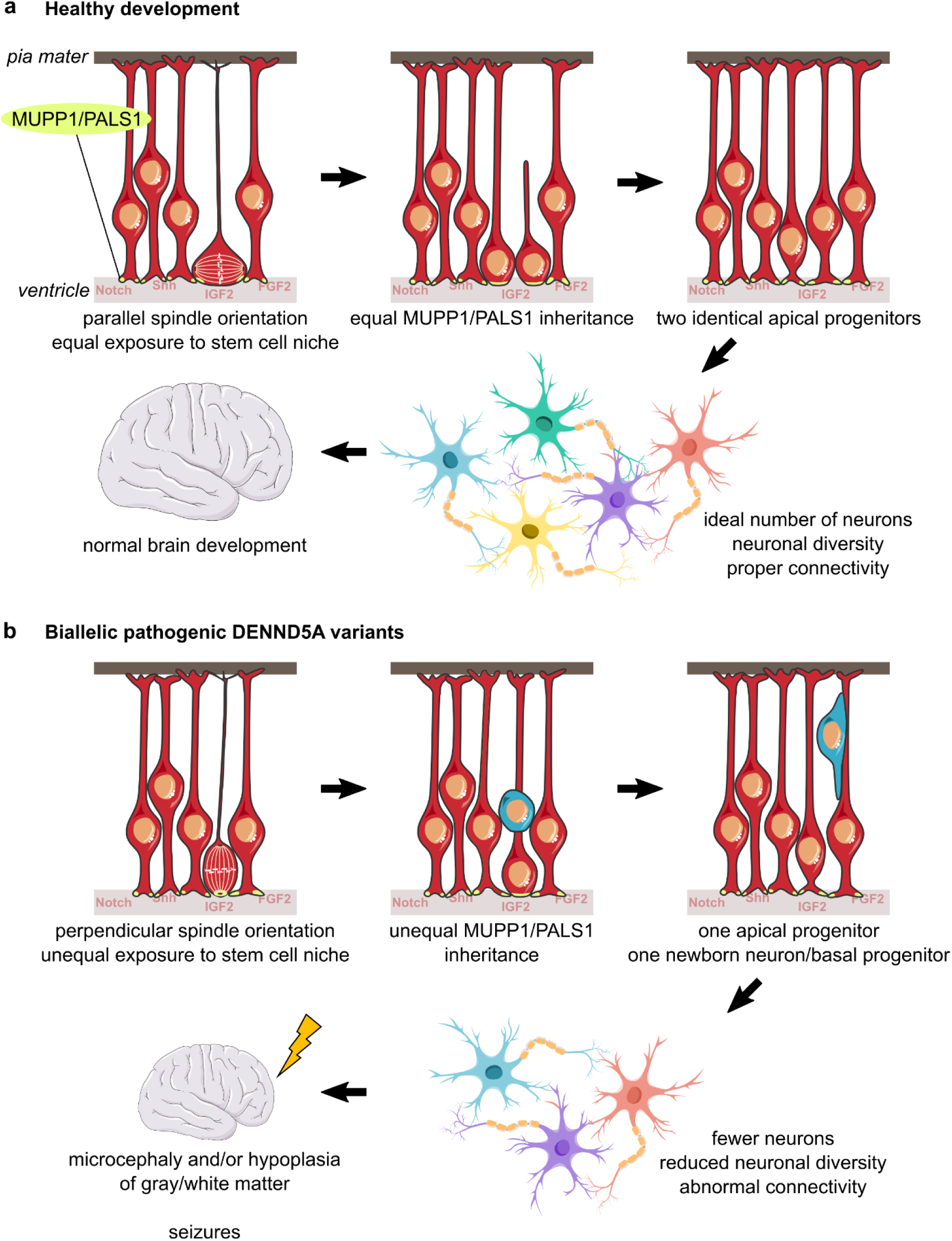
DENND5A-related DEE disease model. a, Under healthy developmental circumstances, apical progenitors are able to obtain a spindle orientation parallel to the apical ventricular surface. This allows both daughter cells to receive equal exposure to the stem and progenitor cell niche as well as inherit equal proportions of apical determinants, such as MUPP1 and PALS1, producing two identical apical progenitors after mitosis. The expansion of the progenitor pool early in brain development allows for an ideal production of neurons from diverse lineages and contributes to healthy brain development. b, In the presence of biallelic pathogenic DENND5A variants, apical progenitors increasingly divide with a spindle angle perpendicular to the ventricular surface. This scenario only allows for one daughter cell to receive signaling molecules from the stem and progenitor cell niche and to inherit apical determinants, and the more basal daughter cell becomes either a basal progenitor or an immature neuron. Increased asymmetric cell division of apical neural progenitors during early development reduces the number of progenitors available for neurogenesis, resulting in a decreased overall number and diversity of neurons that contributes to microcephaly. This may contribute to abnormal neuronal connectivity, resulting in seizures that further adversely affect development, leading to DEE.

---

**Extended Data Figure 1:**
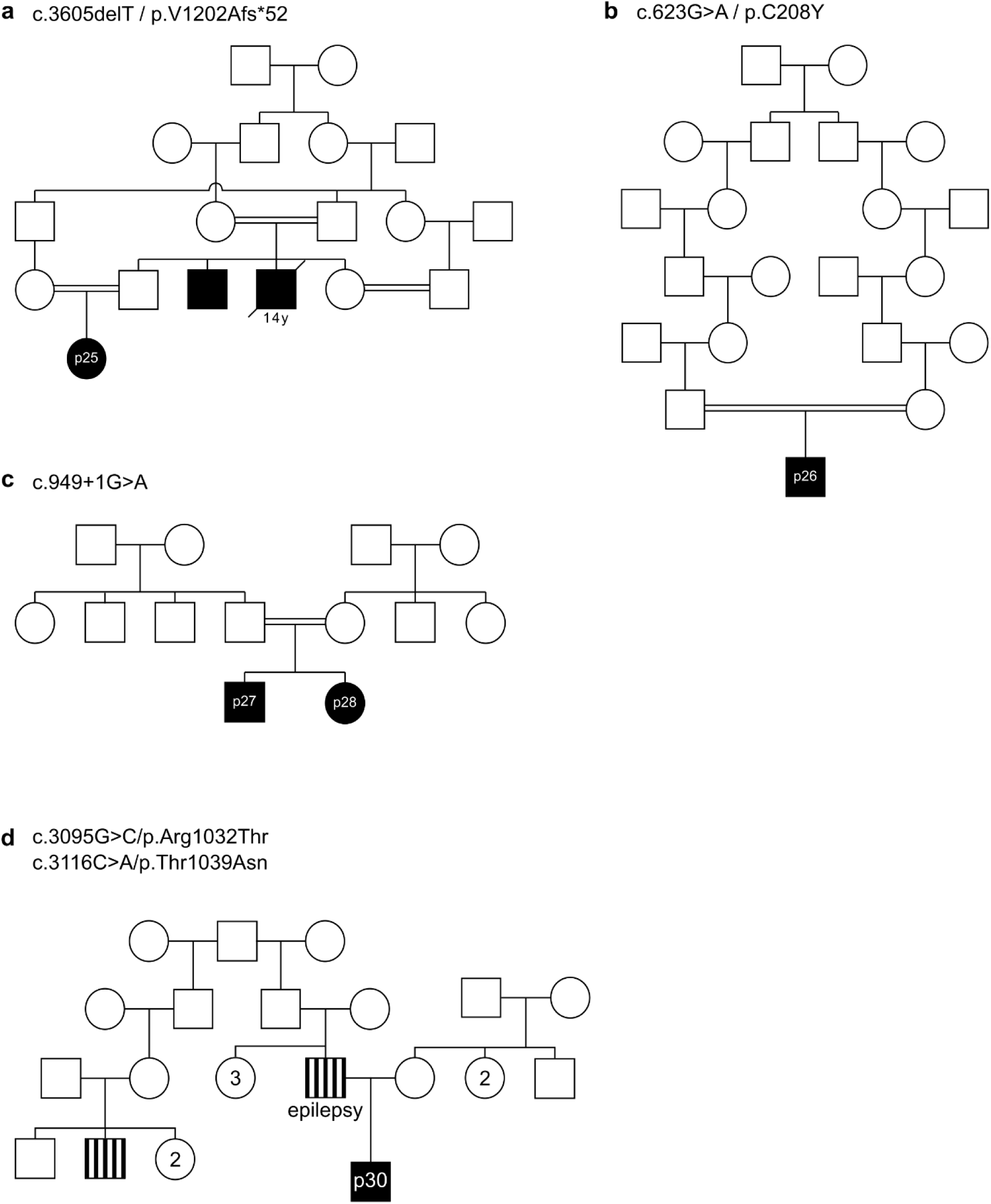
Extended pedigrees of consanguineous families demonstrate pathogenicity of select *DENND5A* variants. Pedigrees indicate affected (colored in) and unaffected (open) individuals in families carrying the variants **a,** c.3605delT/p.V1202Afs*52; **b,** c.623G>A/p.C208Y, **c,** c.949+1G>A, and d, c.3095G>C/p.R1032T and c.3116C>A/p.T1039N. Participants involved in the phenotypic study are indicated by their ID number, and the age at the time of death is indicated for a deceased individual in *(a)*.

**Extended Data Figure 2:**
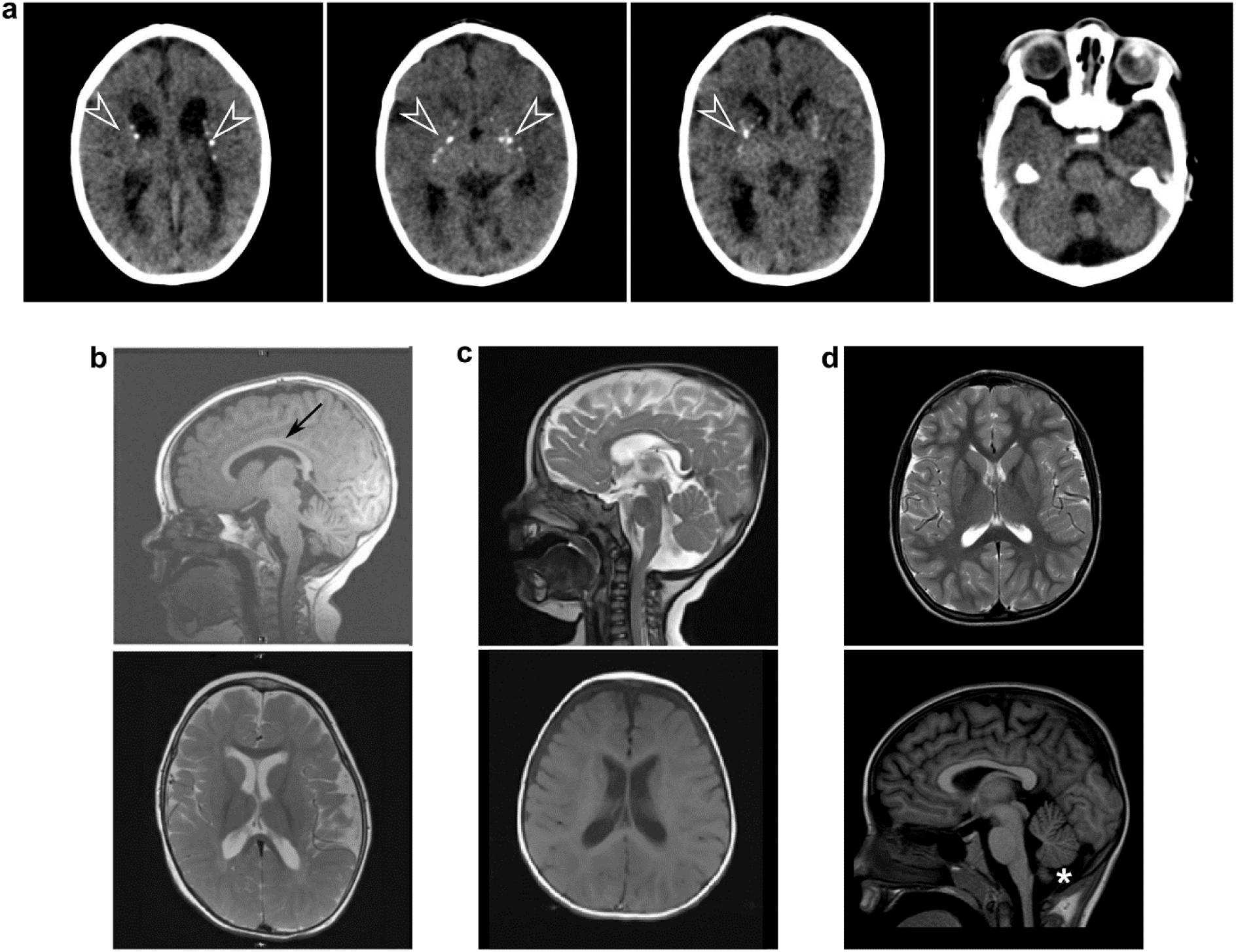
Neuroimaging of other cases with DENND5A-related DEE show varying levels of phenotypic overlap. a, CT from a homozygous individual with the variant p.V1202Afs*52 (participant 25) shows mild cortical volume loss, ventriculomegaly, thin corpus callosum, and lenticulostriate and periventricular calcifications (arrowheads). **b,** MRI from a compound heterozygous individual with variants c.950-20_950-17delTTTT/p.R1078Q (participant 9) shows mild corpus callosum volume loss (arrow). **c,** MRI from a compound heterozygous individual with variants p.R1032T/p.T1039N (participant 30) shows enlarged lateral ventricles. **d,** MRI from a compound heterozygous individual with variants p.K485E/p.R1159W (participant 8) shows a normal MRI with mild inferior cerebellar vermis hypoplasia (asterisk).

**Extended Data Figure 3:**
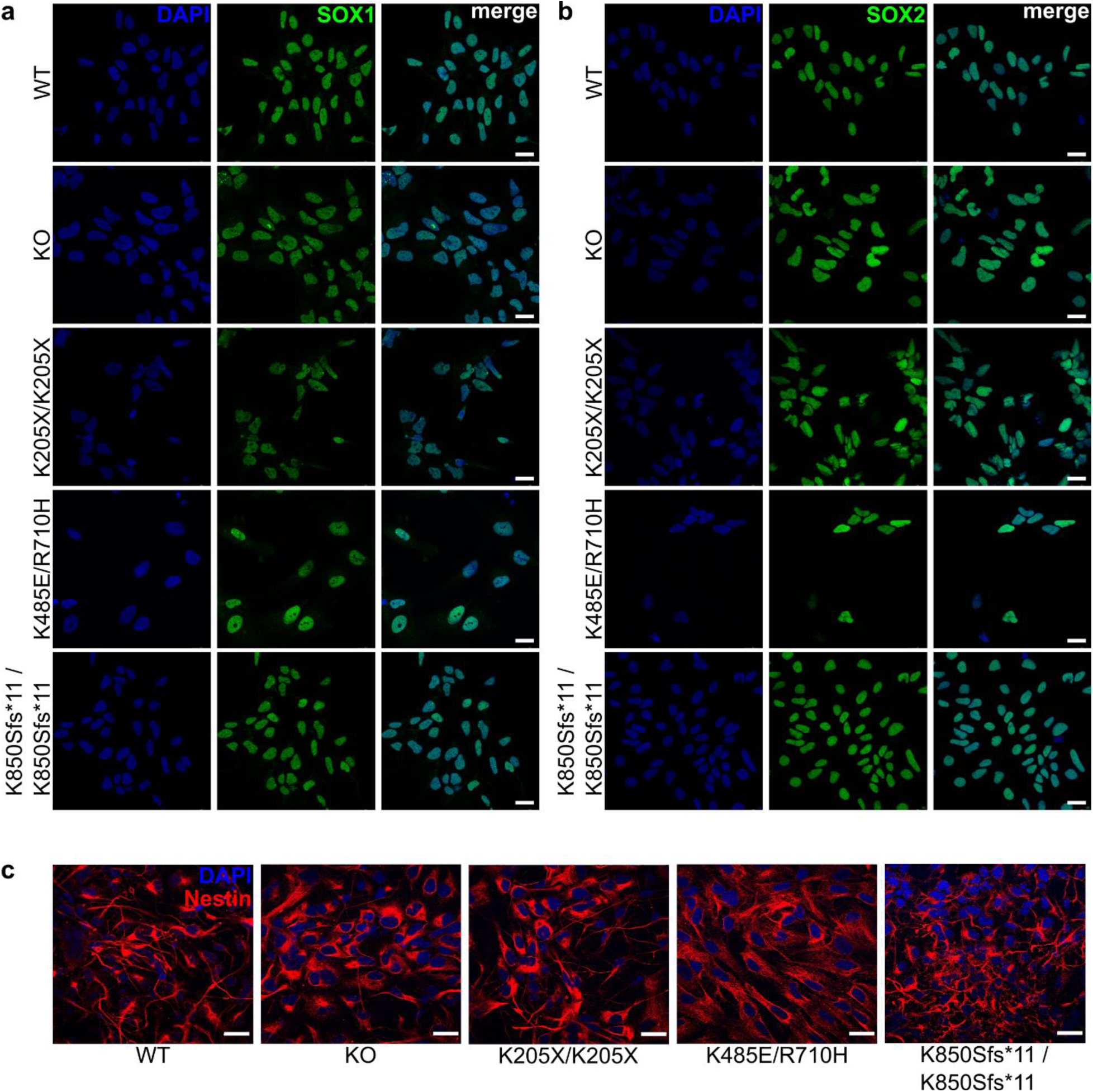
All established NPC lines express neural progenitor-specific markers. iPSCs differentiated into NPCs express **a,** SOX1 (green); **b,** SOX2 (green), and **c,** Nestin (red). Blue = DAPI. Scale bars = 20 µm.

**Extended Data Figure 4:**
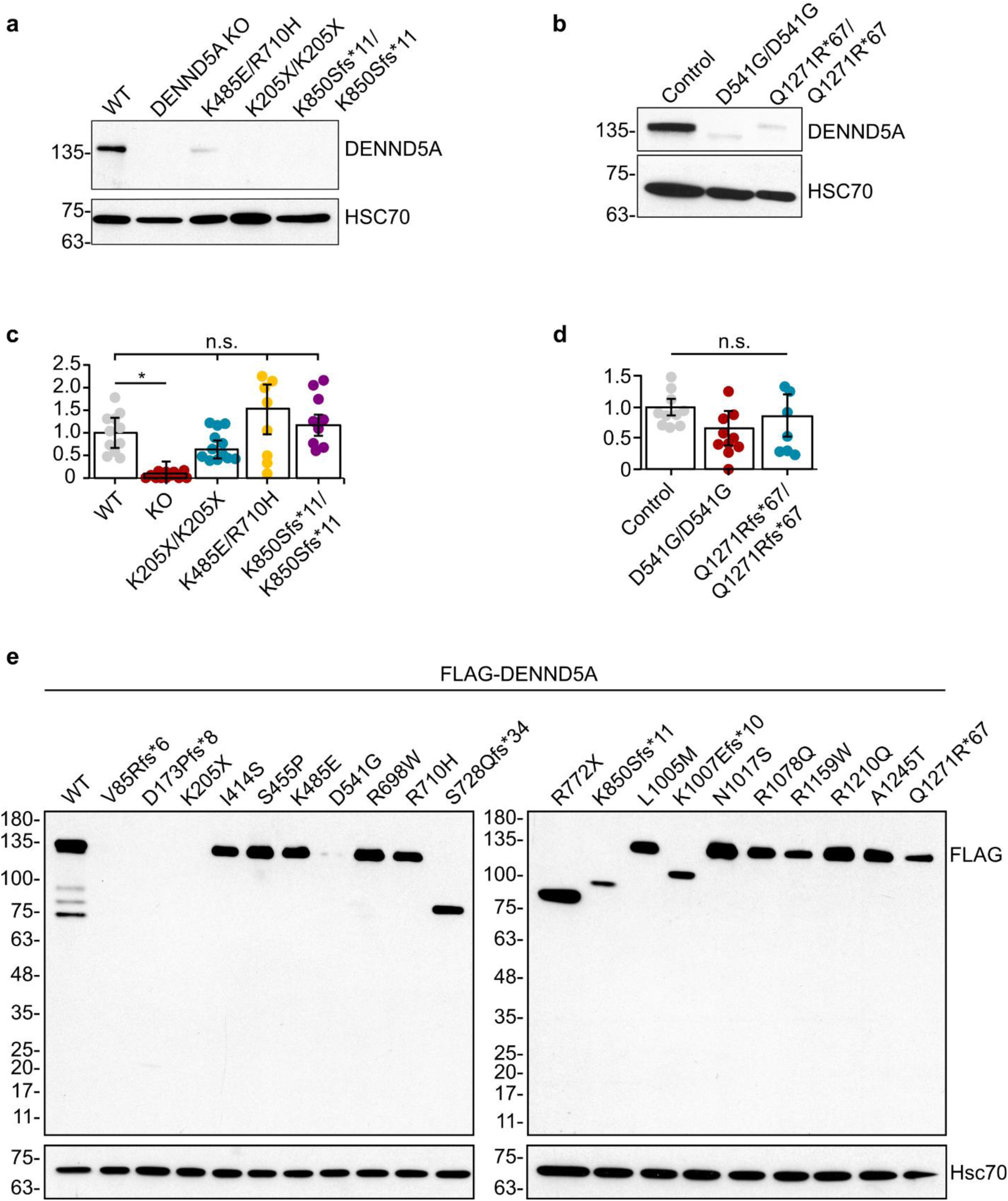
DENND5A expression varies depending on the variant. DENND5A protein expression in **a,** NPCs and **b,** lymphoblasts. Relative *DENND5A* mRNA expression measured by RT-qPCR in **c,** NPCs and **d,** lymphoblasts. Measurements were made with 4 technical replicates on *n* = 3 independent samples. Data are mean ± SEM analyzed via Kruskal-Wallis tests with Bonferroni-corrected pairwise comparisons. **c,** Overexpression of FLAG-DENND5A mutagenized to contain several variants influences protein stability and expression levels in HEK293T.

**Extended Data Figure 5:**
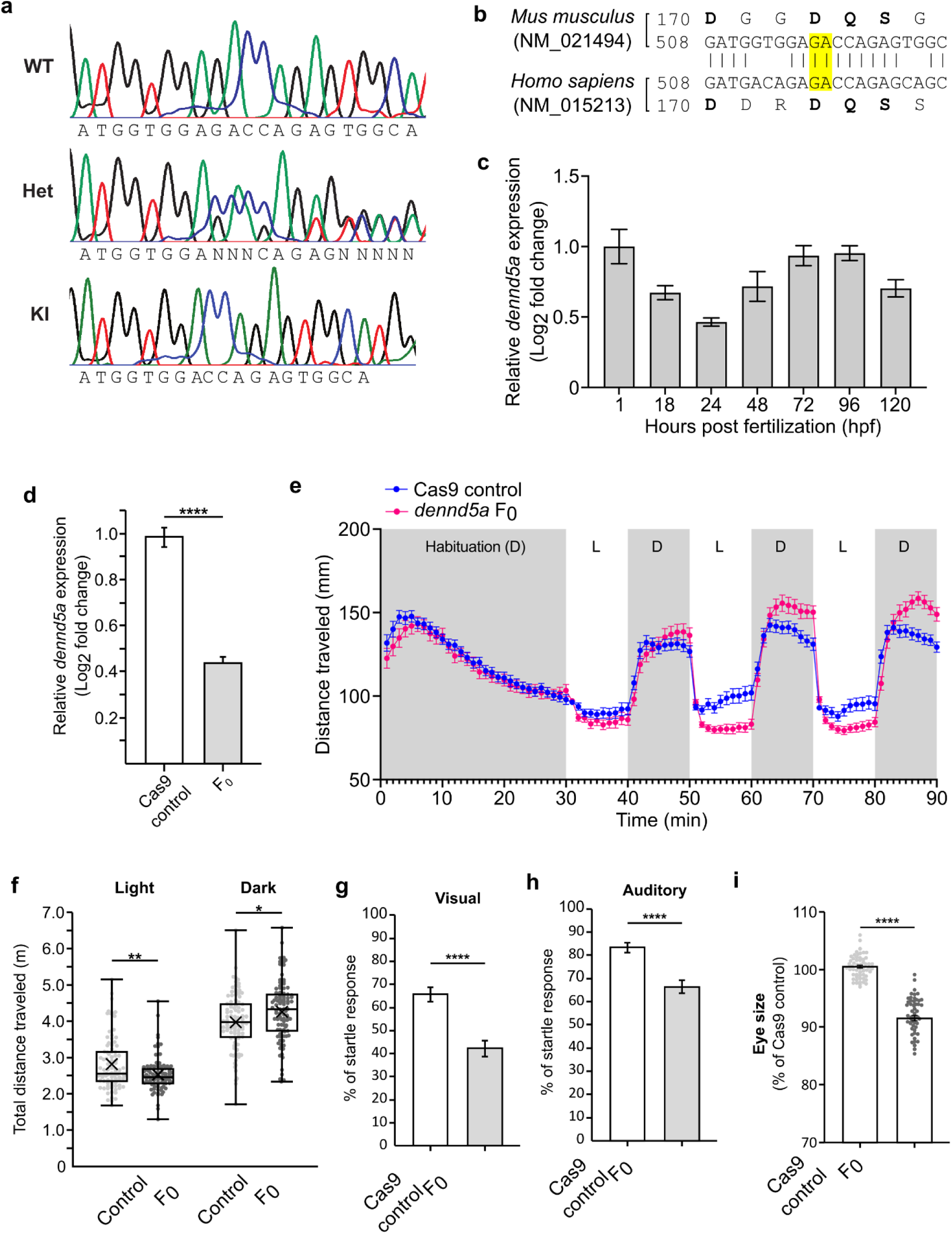
Features of *DENND5A* transgenic animals make them valid models to study *DENND5A*-related DEE. **a,** Sample chromatograms demonstrating DENND5A DNA sequences in WT, heterozygous (Het), and knock-in (KI) mice. **b,** DNA and amino acid sequence alignment between human and mouse DENND5A sequences. Highlighted base pairs indicate bases deleted using CRISPR/Cas9. **c,** The temporal expression of zebrafish *dennd5a* mRNA by RT-qPCR at different developmental stages from experiments performed with biological and technical triplicates. Expression levels were normalized to the *18S* housekeeping gene and compared to 1 hpf embryos. Error bars = mean ± SD. **d,** Expression of *dennd5a* mRNA in Cas9 controls and *dennd5a* F_0_ knockouts detected by RT-qPCR at 5 dpf. Experiments were performed with four biological replicates with technical triplicates. Data are mean ± SEM analyzed via two-tailed student’s *t*-test (*t*(6) = 10.706, *p* < .0001). **e,** Locomotor activities of zebrafish larvae at 5 dpf with n = 96 larvae for each group. Data are mean ± SEM. D = Dark period, L = light period. **f,** Quantification of distance traveled by each larva during the cycles of light or dark periods analyzed via two-tailed Mann-Whitney U test (light; *Z* = −2.81, *p* = .005) and two-tailed student’s *t*-test (dark; *t*(190) = −2.438, *p* = .016). Each dot represents one larva. **g,** Visual startle response in *n* = 143 larvae at 6 dpf. Data are mean ± SEM analyzed via two-tailed Mann-Whitney U test (*Z* = −4.957, *p* < .0001). **h,** Acoustic evoked behavioral response in *n* = 134 larvae at 6 dpf. Data are mean ± SEM analyzed via two-tailed Mann-Whitney U test (*Z* = −4.947, *p* < .0001). **i,** Quantification of eye size in *n* = 60 larvae. Each dot represents one larva. Data are mean ± SEM analyzed via two-tailed Welch’s *t*-test (*t*(96.016) = 17.831, *p* < .0001).

**Extended Data Figure 6:**
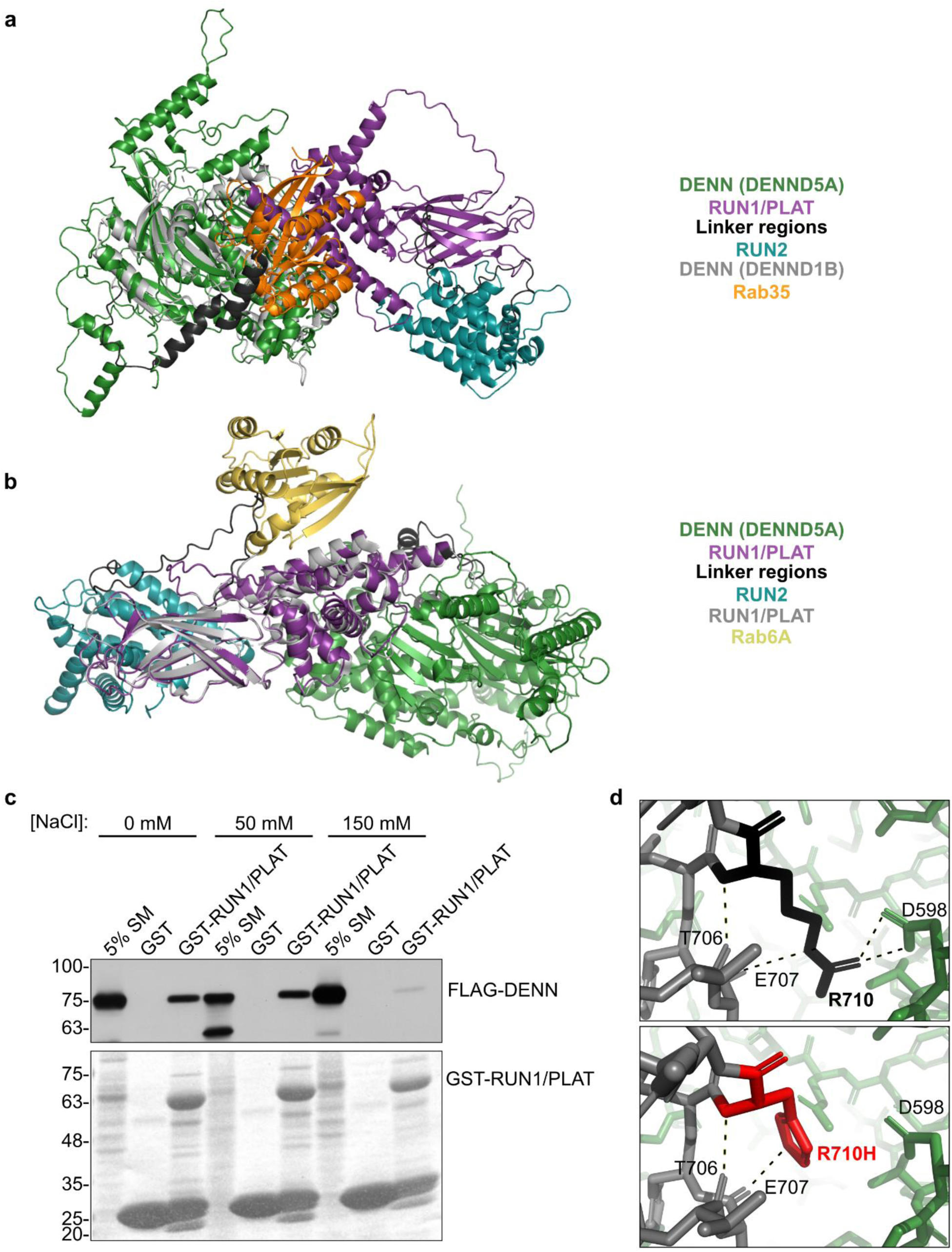
Analysis of the predicted DENND5A structure indicates intramolecular interactions may regulate other protein-protein interactions. **a,** Structural alignment between the predicted DENND5A structure and PDB:3TW8 (gray, yellow) **b,** Structural alignment between the predicted DENND5A structure and PDB:3CWZ (gray, yellow) **c,** Pulldown experiment showing binding capacity between GST-RUN1/PLAT and FLAG-DENN domains of DENND5A under varying NaCl concentrations. **d,** The R710H variant found in the cohort and within the region that interacts with PALS1/MUPP1 results in the removal of two hydrogen bonds with D598 of the DENN domain. Dotted lines indicate hydrogen bonds.

**Extended Data Figure 7:**
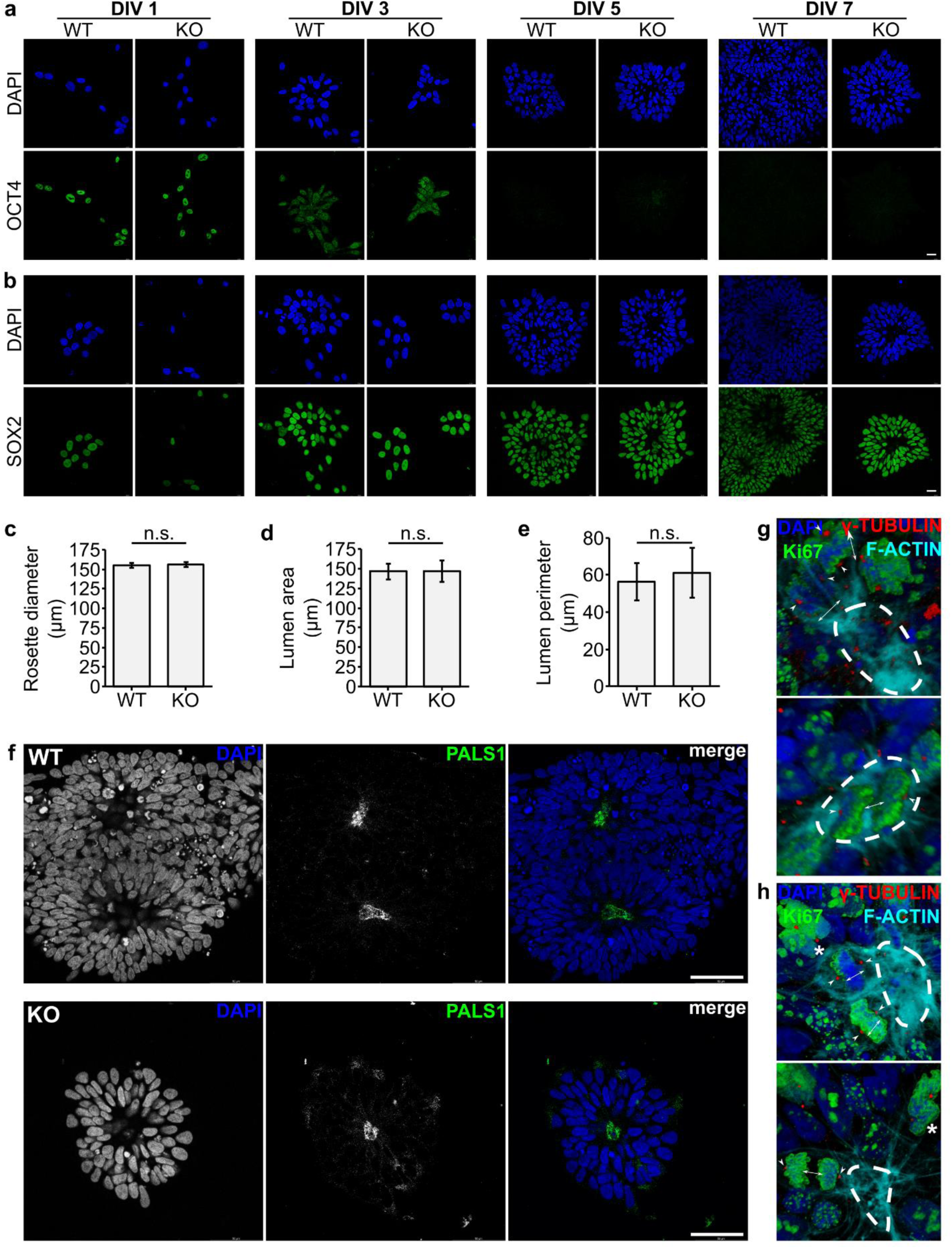
WT and *DENND5A* KO neural rosettes differ in density and cell division properties, but not in marker expression or size. Expression of **a,** OCT4 and **b,** SOX2 during neural rosette development. Blue = DAPI, green = OCT4/SOX2. Scale bars = 20 µm. **c,** Average diameter of individual rosettes. *n* = 159 rosettes were analyzed from 2 independent experiments. Data are mean ± SEM and analyzed via student’s *t*-test. **d,** Average lumen area of rosettes. *n* = 294 rosettes were analyzed from 2 independent experiments. Data are mean ± SEM and analyzed via Mann-Whitney U test. **e,** Average lumen perimeter of rosettes. *n* = 294 rosettes were analyzed from 2 independent experiments. Data are mean ± SEM and analyzed via Mann-Whitney U test. **f,** PALS1 staining (green) shows an apical localization in both WT and KO neural rosettes. Scale bars = 50 µm. **g,** 3D-rendered images of apical progenitors of WT neural rosettes. Blue = DAPI, green = Ki67, red = -tubulin, cyan = F-actin. Arrowheads indicate centrosomes, arrows indicate orientation of cell divisions, dotted lines indicate the lumen. **h,** 3D-rendered images of apical progenitors of KO neural rosettes. Blue = DAPI, green = Ki67, red = -tubulin, cyan = F-actin. Arrowheads indicate centrosomes, arrows indicate orientation of cell divisions, asterisks indicate abnormally condensed chromatin, dotted lines indicate the lumen.

## Acknowledgements

We would like to acknowledge all cohort members in this study, their families, and treating clinicians for their participation. Participant 10 was recruited as part of the German-Jordanian Autosomal Recessive Intellectual Disability project (GeJo-ARID) funded by the German Academic Exchange Service (DAAD) as part of the German-Arab Transformation Program Line4 (Project-ID 57166498). The views expressed are those of the author(s) and do not reflect the official policy of the Department of the Army, the Department of Defense or the U.S. Government. The content is solely the responsibility of the authors and does not necessarily represent the official views of the National Institutes of Health. This work was funded by a Foundation Grant from the Canadian Institutes of Health Research to PSM, and a grant from the Alain and Sandra Bouchard Foundation for Intellectual Disabilities to PSM and TMD. This work was also supported by the King Salman Center for Disability Research through Research Group no RG-2022-010 (FSA). This work was also performed under the Care4Rare Canada Consortium funded by Genome Canada and the Ontario Genomics Institute (OGI-147), the Canadian Institutes of Health Research, Ontario Research Fund, Genome Alberta, Genome British Columbia, Genome Quebec, and Children’s Hospital of Eastern Ontario Foundation (DC, ESYG, MO). Research reported in this manuscript was also supported by the NIH Common Fund, through the Office of Strategic Coordination/Office of the NIH Director under Award Number(s) [U01HG007672, U01HG007943] (HC) and [NS105078, HG011758] (JRL).

## Author Contributions

EBan designed, performed, and analyzed most experiments and wrote the manuscript. VFr performed mouse immunohistochemistry, coordinated induced seizure experiments and mouse MRIs, and maintained the mouse colony. SJL performed and analyzed zebrafish experiments in Fig. 4 and Extended Data Fig. 5. FK established the mouse colony, performed the western blot in Fig. 3a, and coordinated mouse MRIs. VFo processed and analyzed mouse MRIs. ML performed the induced seizure experiments. CH established the iPSC lines. GK performed RT-qPCR on mouse brain tissue for Fig. 3b. MT performed the mouse MRIs. AB analyzed mouse MRIs and provided experimental input. FSA generated and provided lymphoblast cell lines and contributed two clinical cases. SH and SK established iPSC lines from participants 2 and 10. RM identified many cohort members and provided pedigrees for Extended Data Fig. 1. RAK, FSA, LA, MBab, MBah, BB, EBar, LB, MBas, MBer, DB, RB, Mbu, MC-D, DCa, HC, DCu, SE, MAE, HGES, TF, HKG, JGG, LG, ESYG, VKG, TBH, MOH, TLH, JSH, AH, HH, KH, SH, EGK, GCK, AK, HL, JRL, EJM, AM, DM, JN-F, HN, DN, BO-J, MO, DP, LGS, CS, LS, VS, RCS, VMS, MSZ, DZ, and CZ completed phenotypic questionnaires and contributed to clinical phenotyping genetic analysis. KM acted as a liaison to connect EBan with clinicans that submitted variants on ClinVar. RP analyzed clinical exomes of GeneDx-tested cases. BAM identified the initial cases. UDN is associated with the contribution of data from participant 9. KH and CP provided technical aid to F_0_ zebrafish experiments. J-FT identified the intramolecular interface in Fig. 4 and supervised all structural analyses. TMD supervised iPSC quality control and cell line production. ZG supervised genetic analysis. MA supervised induced seizure experiments. CA analyzed participant MRIs and CTs. GKV supervised zebrafish experiments. RM contributed many clinical cases and connected us with CA, GKV, SJL, KH, and CP. All authors reviewed the manuscript for accuracy and edited wording or data presentation according to their clinical, molecular, structural, genetic, or biological expertise. PSM supervised the study, designed experiments, secured funding, and wrote the manuscript.

## Additional Information

Supplementary Information is available for this paper.

Correspondence and requests for materials should be addressed to PSM. Reprints and permissions information is available at www.nature.com/reprints.

## Methods

### Participant Recruitment

All materials and methods for participant recruitment and clinical data collection was approved by the McGill University Health Centre research ethics board (study 2021-6324) and the McGill Faculty of Medicine and Health Sciences institutional review board (study A12-M66-21B). All participants were recruited based on the presence of at least one variant in the gene *DENND5A*. Recruitment occurred through clinicians directly contacting our laboratory (16 participants) based on a previous publication about DENND5A^1^ (4 participants), through GeneMatcher^3^ or ClinVar^4^ genetic databases (5), through contacting corresponding authors of other publications that briefly described patients with DENND5A variants^2^ (2), and through word-of-mouth between collaborators of our study (3). Sex and gender were not considered in study design, and the research findings do not apply to only one sex. Pathogenic variants, likely pathogenic variants, and variants of unknown clinical significance were all eligible for inclusion in the study. Six individuals were ineligible and were excluded from analysis: 2 participants were heterozygous for a *DENND5A* variant, 1 was excluded due to death occurring prior to clinical data collection^1^, and 3 were excluded because the questionnaires were not returned. Recruitment spanned approximately four years.

### Phenotypic data collection and analysis

Clinicians with patients harboring biallelic *DENND5A* variants completed an anonymized phenotypic questionnaire based on their patient’s most recent clinic visit. Available anonymized MRIs, CTs, and/or or official reports were contributed if the patient underwent neuroimaging for clinical purposes. Participants were assigned a numerical ID in the order in which their questionnaires were received. For intronic variant analysis, molecular consequences were predicted using Ensembl’s Variant Effect Predictor. ^5^ For those whose raw MRI or CT data were provided, an independent neuroradiologist re-analyzed the scans and completed the “Brain” section of the questionnaire without viewing the original submitted questionnaires. If responses to an item differed between the original clinician and the independent radiologist, the independent radiologist’s response was used for analysis. Data are missing if the presence of a phenotype is officially unknown. The most frequently reported phenotypes (present in >50% of the cohort) were arranged into a Venn diagram using an online tool (https://www.meta-chart.com) to illustrate the degree of phenotypic overlap within and between participants. For occipitofrontal circumference (OFC), if percentiles were not given directly from clinicians, percentile values were derived from the age- and sex-appropriate Word Health Organization tables (https://www.who.int/tools/child-growth-standards/standards/head-circumference-for-age). The OFC percentile for one person whose measurements were taken when they were above 5 years old was derived from tables published in Adela Chirita-Emandi et al^6^. For calculating central tendency statistics, OFC percentiles given as a range (e.g. < 3 or < 1) were assigned a conservative numerical estimate (e.g. 2.9 for < 3, 0.9 for < 1).

### Establishment of cell lines

The control induced pluripotent stem cell (iPSC) line AIW001-02 was derived from peripheral blood mononuclear cells of a healthy female donor (Caucasian, 48 years old). The AIW001-02 cell line was generated by using the CytoTune™-iPS 2.0 Sendai Reprogramming Kit (iPSQuebec Platform, Laval University). For knockout of human *DENND5A*, guide RNAs (gRNAs) were designed using an online tool (https://benchling.com). Both gRNA target sites are on *DENND5A* exon 4. Synthesized gRNAs were ordered from SYNTHEGO and transfection was performed following the manufacturer’s protocol. Single cell colonies were picked and amplified. Genomic DNA from the colonies was extracted with QuickExtract (Lucigen) and PCR was performed using Q5 High-Fidelity DNA Polymerase according to the manufacturer’s protocol (F: GAGGATCGCCAGTGAGTGTT; R: CCCCGAGCAGTTCAAAAACC). A 238 base pair deletion was confirmed by Sanger sequencing.

Human fibroblasts from DENND5A cohort members were obtained by skin biopsy (participants 2 and 10) and renal epithelial cells (participant 3) from a urine sample. Cells were reprogrammed to iPSCs by electroporation with episomal plasmids (pCXLE-hUL, pCXLE-hSK, and pCXLE-hOCT4) as previously described^7^. Generated iPSCs were functionally and genomically validated according to Hauser and Erzler^8^.

Lymphoblasts were obtained from a healthy individual (control line) and two homozygous individuals (participants 4 and 5). Cells were immortalized through use of the Epstein-Barr virus and generated in the lab of Dr. Fowzan Alkuraya.

### Cell culture

iPSCs were cultured on hESC-qualified Corning Matrigel-coated tissue culture dishes in either TeSR-E8 medium (all patient-derived iPSC lines; STEMCELL Technologies) or mTeSR1 medium (AIW001-02 WT and *DENND5A* KO; STEMCELL Technologies) with daily medium changes and mechanical removal of differentiated cells. Cells were passaged using the ReLeSR Passaging Reagent (STEMCELL Technologies) once cultures reached approximately 70% confluency.

iPSCs were differentiated to neural progenitor cells (NPCs) using the STEMdiff SMADi Neural Induction Kit (STEMCELL Technologies) with daily medium changes. Induced cultures were passaged using Accumax (Millipore Sigma) once cells reached 90-95% confluency, approximately once per week. After a two week induction period, NPCs were maintained in STEMdiff Neural Progenitor Medium (STEMCELL Technologies) on poly L ornithine (PLO)- and laminin-coated plates and passaged using Accumax once cultures reached 80-95% confluency, approximately once per week. Experiments examining β-III tubulin expression examined established NPC lines after one passage post-neural induction; all other experiments were performed using cells at passages 2-4.

Control and patient-derived Epstein-Barr virus-induced lymphoblastoid cell lines were obtained from the laboratory of Dr. Alkuraya. Cells were cultured in suspension in RPMI 1640 medium (Gibco) supplemented with 15% fetal bovine serum (Wisent), 1% penicillin-streptomycin (Wisent), and 1% L-glutamine (Wisent). Cells were split 1:4 once confluency reached approximately 1 x 10^6^ cells/ml.

For biochemical studies, HEK293-T cells were cultured in DMEM high glucose (Fisher) supplemented with 10% bovine calf serum (Fisher), 1% L-glutamine (Wisent), and 1% penicillin-streptomycin (Wisent).

Because this autosomal recessive disease appears to affect males and females approximately equally, sex was not considered in study design when conducting *in vitro* experiments.

### Plasmid cloning

*DENND5A* cDNA (Origene, SC121400) was cloned into the pCMV-tag2B vector to generate FLAG-DENND5A. GFP-DENND5A was made via subcloning DENND5A into the pEGFP-C1 vector. Patient variants and targeted residues for biochemical studies were introduced using the QuikChange Lightning site-directed mutagenesis kit (Agilent) following the manufacturer’s protocol. FLAG-DENND5A DENN domain was made by subcloning aa1-680 of DENND5A into the pCMV-tag2B vector. GST-aa700-720 was made via oligo annealing followed by ligation into a pGEX-4T1 vector with a modified multiple cloning site (MCS). GST-RUN1/PLAT was created by subcloning DENND5A aa707-1090 into the pGEX-6P1 vector. MUPP1 (MPDZ) was obtained from the Harvard Medical School plasmid collection (HsCD00352820). Untagged PALS1 (MPP5) in pDONR223 was obtained from Addgene (#23447) and subcloned into a pCMV3-C-FLAG vector to generate PALS1-FLAG. The vector backbone from PALS1-FLAG was then isolated and modified to include a custom MCS via oligo annealing and ligation in order to create restriction sites suitable for subcloning MUPP1 into the vector. MUPP1 was subcloned into this modified vector to create MUPP1-FLAG. All constructs were confirmed by Sanger sequencing.

### RT-qPCR

RNA was extracted from NPCs and lymphocytes using the RNeasy kit (Qiagen) followed by cDNA synthesis using iScript Reverse Transcription Supermix for RT-qPCR (Bio-Rad). RT-qPCR was performed using SsoFast EvaGreen Supermix (Bio-Rad) with primers targeting human *DENND5A* (F: CTAAAGCCAGGGATGGTGCC; R: TTTCGGCATACATAGCATTCCT) and *TBP* (F: TGCACAGGAGCCAAGAGTGAA; R: CACATCACAGCTCCCCACCA). *DENND5A* levels were normalized to *TBP* levels and AIW001-02 WT NPCs or control lymphocytes.

RNA from mouse brain tissue was extracted using RNeasy Lipid Tissue kit (Qiagen), followed by cDNA synthesis and RT-qPCR performed with the same reagents as above using primers specific to the mouse genome for *DENND5A* (F: CAGTCGCTTCGCCGACTAT; R: GCACCATCCCTGGCTTTAGAA) and *GAPDH* (F: ACTCCACTCACGGCAAATTC; R: CCAGTAGACTCCACGACATACT). *DENND5A* levels were normalized to *GAPDH* levels in WT mice.

RNA was extracted from zebrafish samples using TRIzol Reagent (Thermo Fisher Scientific) and purified with the RNA Clean and Concentrator-5 kit (Zymo Research) according to the manufacturer’s guidelines. cDNA was synthesized using iScript Reverse Transcription Supermix (Bio-Rad). RT-qPCR was performed on the Light Cycler 96 System (Roche) using specific primers for zebrafish *dennd5a* (F: TTGGCGAGTGCAAATGAAGG; R: GTTTGCTGGTCACCTCCTCA) along with SYBR Green Supermix (Thermo Fisher Scientific). *dennd5a* expression levels were normalized to the *18S* gene and the 1 hpf stage (in temporal *dennd5a* expression analysis) or to Cas9-injected control animals (to analyze *dennd5a* expression in control versus F_0_ KO animals).

### Neural rosette formation assay

iPSCs were gently dissociated into a single cell suspension and plated at low density (20,000 cells per well of a 24 well plate) onto PLO/Laminin-coated coverslips in neural induction media with SMAD inhibitor (STEMCELL Technologies #08581) containing 10 µM Y-27632 on the day of plating. Medium was changed daily and cells were fixed after 1, 3, 5, and 7 days in culture. Rosettes were stained and z-stack images in 0.5 µm increments were taken using the Leica SP8 confocal microscope. For lumen size analysis, z-plane images with the largest visible lumens were used for quantification followed by two-tailed student’s *t*-tests. For rosette diameter analysis, z-plane images with the widest phalloidin staining were used for quantification followed by a two-tailed student’s *t*-test. To analyze dividing cells within the rosettes, cells in metaphase, anaphase, and telophase were used for quantification. Mitotic spindle angles were measured as in Chilov et al., 2011^9^ using F-actin as a marker of the apical surface and quantified using ImageJ.

### Immunocytochemistry

Samples were fixed in 4% paraformaldehyde for 20 minutes at room temperature. Cells were permeabilized for 5 minutes in 0.1% Triton-X 100 and then blocked in 5% BSA and 0.01% Triton-X 100 in PBS for 30 minutes at room temperature, followed by overnight incubation at 4°C with the following antibodies: OCT4 (ab19857, 1 µg/ml), SOX1 (Invitrogen MA5-32447, 1:200), SOX2 (Abcam ab92494, 1:1000), Nestin (Invitrogen, MA1-110), β-III tubulin (Abcam ab52623, 0.1 µg/ml), Ki67 (Abcam ab15580, 0.5 µg/ml), γ-tubulin (Sigma-Aldrich T6557, 1:500), and Pals1 (Santa Cruz Biotechnology sc-365411, 1:350). After primary antibody incubation, samples were washed twice with PBS then incubated with Alexa-conjugated secondary antibodies at 1:500 dilution and phalloidin at 1:1000 dilution for 1 hour at room temperature. Cells were then washed twice in PBS, incubated with DAPI at 1:5000 dilution for 10 minutes, washed twice in PBS again, and mounted onto glass slides.

### Endogenous and transfected protein expression

NPCs and lymphoblasts were harvested in lysis buffer (20 mM HEPES pH 7.4, 100 mM NaCl, 0.83 mM benzamidine, 0.5 µg/ml aprotinin, 0.5 g/ml leupeptin, 0.23 mM phenylmethylsulfonyl fluoride) containing 1X LSB. Mouse brains were homogenized in lysis buffer and incubated with 1% Triton X-100 for 15 minutes at 4°C, followed by centrifugation at 239,000 x *g* for 15 minutes at 4°C. For FLAG-DENND5A expression, plasmid DNA was transfected into HEK293T cells using the calcium phosphate method and cells were harvested after 24 hours in lysis buffer containing 1X LSB. Equal protein aliquots were loaded onto an SDS-PAGE gel and analyzed via Western blot using primary antibodies against DENND5A (ThermoFisher #702789, 2.5 µg/ml), FLAG (clone M2, Sigma-Aldrich, 1:10,000), Hsc70 (clone 1B5, StressGen Biotechnologies Corp, 1:1:10,000), and β-actin (Clone C4, Sigma-Aldrich MAB1501R, 1:1000).

### Pulldown experiments

500 mL flasks of bacteria were induced overnight at RT to express GST-tagged proteins using 500 µM IPTG when the optical density of the cultures at 600 nm reached 0.6. Bacteria were pelleted and resuspended in PBS + protease inhibitors pH 7.4. Resuspended bacterial cells were then sonicated 3 times for 5 seconds at 70% amplitude, followed by incubation with 1% Triton X-100 for 15 minutes. Bacterial cell lysate was then spun for 5 minutes at 4°C at 11,952 x *g*. The supernatant was incubated with glutathione Sepharose beads pre-washed 3 times in PBS for one hour at 4°C, and beads were then briefly spun down and washed 3 times in PBS + protease inhibitors to purify GST fusion proteins. The concentration of fusion proteins was determined by running on an SDS-PAGE gel accompanied by a BSA standard curve followed by Coomassie Brilliant Blue staining. Cell lysates were then harvested for incubation with GST fusion proteins.

For pulldown experiments with overexpressed proteins, HEK293-T cells were transfected using the calcium phosphate method with the appropriate plasmids and harvested the next day in lysis buffer. Cells were then sonicated once for 10 seconds at 20% amplitude, incubated in 1% Triton X-100 for 15 minutes at 4°C, and spun at 21 x *g* and 4°C for 15 minutes in a tabletop centrifuge. The concentration of HEK293T supernatants were analyzed via a Bradford assay, and 1 mg/ml was incubated with 20 µg (GST-aa700-720) or 100 µg (GST-RUN1/PLAT) of fusion proteins for one hour at 4°C. Following incubation, beads were washed 3 times in buffer containing 1% Triton, eluted in 1X LSB, loaded onto an SDS-PAGE gel, and analyzed via Western blot using primary antibodies against FLAG (clone M2, Sigma-Aldrich, 1:10,000).

### Mass Spectrometry

For the initial protein-protein interaction screen followed by mass spectrometry, 4 E18 rat brains were homogenized on ice in 1 ml lysis buffer per brain (20 mM HEPES pH 7.4, 150 mM NaCl, 0.83 mM benzamidine, 0.5 μg/ml aprotinin, 0.5 μg/ml leupeptin, and 0.23 mM phenylmethylsulfonyl fluoride) with 10 strokes using a Caframo homogenizer at 1200 rpm. Tissue homogenate was then sonicated once for 10 seconds at 20% amplitude, incubated in 1% Triton X-100 for 15 minutes at 4°C, and spun at 239,000 x *g* and 4°C for 15 minutes. Supernatant concentration was determined via a Bradford assay and 1 mg/ml was incubated with 50 µg GST-fusion protein overnight at 4°C. Following incubation, beads were washed 3 times in lysis buffer containing 1% Triton, then eluted in 1X Lammeli sample buffer (LSB).

For each sample, proteins were loaded onto a single stacking gel band to remove lipids, detergents, and salts. The gel band was reduced with DTT, alkylated with iodoacetic acid, and digested with Trypsin. Extracted peptides were re-solubilized in 0.1% aqueous formic acid and loaded onto a Thermo Acclaim Pepmap (Thermo, 75 µM ID X 2cm C18 3 µM beads) precolumn and then onto an Acclaim Pepmap Easyspray (Thermo, 75 µM X 15cm with 2 µM C18 beads) analytical column separation using a Dionex Ultimate 3000 uHPLC at 250 nl/min with a gradient of 2-35% organic (0.1% formic acid in acetonitrile) over 2 hours. Peptides were analyzed using a Thermo Orbitrap Fusion mass spectrometer operating at 120,000 resolution ( FWHM in MS1) with HCD sequencing (15,000 resolution) at top speed for all peptides with a charge of 2+ or greater. The raw data were converted into *.mgf format (Mascot generic format) for searching using the Mascot 2.6.2 search engine (Matrix Science) against all rat protein sequences (Uniprot 2017). Search parameters for peptides > 5 residues were +/− 5 ppm on the parent ion and 0.1 amu on fragment ions. A fixed modification for carboxymethyl-Cysteine was used along with variable modifications of Oxidation (methionine) and deamination (asparagine/glutamine). At 99.0% protein and 95% peptide confidence, 1077 proteins (34,443 spectra) were identified using 1 peptide (0.0% peptide FDR and 0.40% protein FDR). The database search results were loaded onto Scaffold Q+ Scaffold_4.8.6 (Proteome Sciences) for statistical treatment and data visualization.

### Co-immunoprecipitation experiments

For co-immunoprecipitation experiments, HEK293-T cells were transfected using the calcium phosphate method, harvested in lysis buffer, and sonicated once for 10 seconds at 20% amplitude, followed by incubation in 0.5% Triton X-100 for 15 minutes at 4°C. Cell lysate was then spun at 21 x *g* and 4°C for 15 minutes in a tabletop centrifuge, and 1 mg/ml of the resulting supernatant was incubated for one hour at 4°C with 25 µl ChromoTek GFP-Trap Agarose magnetic beads pre-equilibrated 3 times in lysis buffer without Triton X-100. Beads were then washed 3 times in lysis buffer containing 0.05% Triton and then eluted in 1X LSB for SDS-PAGE analysis and analyzed via Western blot using primary antibodies against FLAG (clone M2, Sigma-Aldrich, 1:10,000) and GFP (Invitrogen Cat# A-6455, 1:20,000).

### Animal care and selection

All mouse care and experiments in the study were approved by the Montreal Neurological Institute Animal Care Committee in accordance with guidelines set by the Canadian Council on Animal Care under ethical protocol number 5734. The experimental unit for this study is a single animal. Apart from selecting animals based on *DENND5A* genotype, no exclusion criteria were set for the experiments. Sex was not considered in study design because there is an approximately equal distribution of males and females in the cohort, so both male and female animals were used.

Crispant zebrafish experiments used wild-type strain NHGRI-1^10^ according to the protocol approved by the Institutional Animal Care Committee (IACUC) of Oklahoma Medical Research Foundation (22-18). All animals were raised and maintained in an Association for Assessment and Accreditation of Laboratory Animal Care (AAALAC) accredited facility under standard conditions.

### Establishment of transgenic animal models

KI mice were generated by the McGill Integrated Core for Animal Modeling. Two silent mutations were introduced in L168 and L169 (CTTGCT –> TTAGCA) as well as a deletion of 2 bp in G172 to introduce a frameshift and premature stop codon in exon 4 of the *DENND5A* mouse gene. Briefly, custom sgRNAs (Synthego), Cas9 protein (IDT, Cat#1081058) and ssODN (ultramer, IDT) were microinjected into the pronucleus of C57BL/6N mouse zygotes with concentrations of 50:50:30 ng/µl respectively. Embryos were subsequently implanted in CD-1 pseudopregnant surrogate mothers according to standard procedures approved by the McGill University Animal Care Committee. Founder pups (F0) were genotyped for evidence of a deletion of 2 bp in G172 and mated to wild-type C57BL/6N (Charles River) mice for three generations. The colony was maintained by sibling mating and by crosses to C57BL/6N mice every third generation. All genomic sequencing was performed using the Big Dye Terminator Ready Reaction Mix (ABI, Carlsbad, CA, USA) at the McGill and Genome Quebec Innovation Center (Primers: ACAAGGAATGCTCTCACTGC, CACACTCCGACATGCCTTCAT [417 bp]). Obtained sequences were analyzed using an online tool (https://benchling.com).

Previously described methods were used to generate *dennd5a* KO zebrafish in crispant experiments^11,12^ using single-guide RNAs (sgRNAs) designed using the CRISPOR tool targeting three *dennd5a* sites (GGTGTTTGAGCTGCTAGGGC, GGGCTGATCTGACAGGAAGG, AGATGGGCCATGATAACTCA) and synthesized by in vitro transcription. Embryos at the one-cell stage were injected with a mixture containing Cas9 protein with (F_0_ knockouts) or without (control) sgRNAs.

### 4-aminopyridine induced seizure assay

6 month old mice (*M* = 183.9 days, *SD* = 1.0) were injected with the K+ channel blocker 4-aminopyridine (8 mg/kg, i.p.) (Sigma-Aldrich, Canada) to induce seizures. If no seizures were observed after 30 min, they were re-injected with a half-dose of 4-aminopyrdine (4 mg/kg, i.p.). Animals that showed no seizures after the second dose were excluded from further analysis. Seizures were identified based on behavioral symptoms such as myoclonic activity of rear and forelimbs that evolved to rearing and loss of balance. The latency (min) from the time of the last 4-aminopyridine injection and seizure onset was calculated.

### Zebrafish morphological and behavioral phenotyping

Morphological phenotyping was conducted by randomly selecting Cas9-injected control and F_0_ KO animals at 5 days post fertilization (dpf) and anesthetizing them with Tricaine/MS-222 (Sigma-Aldrich). The animals were positioned in 2% methylcellulose (Sigma Aldrich) under a stereomicroscope and imaged using a high-definition Nikon DS-Fi2 camera mounted on a Nikon SMZ18 stereomicroscope. Head and eye sizes were measured from scale-calibrated images using ImageJ.

All behavioral tests were conducted at room temperature. Acoustically evoked behavioral response tests were carried out using Zebrabox behavior chambers (Viewpoint Life Sciences) as previously described^12^ involving the percentage of responses for 12 stimuli per larva. The visual startle response test was performed using the DanioVision system running EthoVision XT software (Noldus Information Technology, Leesburg), as previously reported^12,13^ based on the number of responses for five stimuli per larva. For locomotor behavior recording during light/dark transitions, 4 dpf larvae were transferred to a 96-well plate, with one larva per well in 150 µL of embryo water. The following day, the plate was placed in a Noldus chamber, and the DanioVision system running EthoVision XT software was used to record locomotion activity. The 5 dpf larvae underwent a 30-minute habituation period in the light, followed by three cycles of 10-minute light and 10-minute dark transitions. The locomotion activity of the larvae was recorded as the distance traveled in millimeters (mm) per minute.

### Whole-mount in situ hybridization (WISH)

WISH was performed on zebrafish embryos using a previously described method^14^ using a 673-bp amplicon of zebrafish dennd5a cDNA produced via PCR (Primers: T3-dennd5a_F: gaattgaattaaccctcactaaagggCGAGTGCAAATGAAGGTGAG; T7-dennd5a_R: gaattgtaatacgactcactatagggGGTCTCTGACAGATCACTGG).

### Immunohistochemistry

Mouse brain sections from 124 day old mice were baked overnight at 60°C in a conventional oven. Samples were then deparaffinized and rehydrated in a series of xylene and ethanol washes, followed by antigen retrieval using citrate buffer (pH 6.0) for 10 minutes at 120°C in a decloaking chamber (Biocare Medical). Slides were then rinsed IHC buffer (PBS + 0.05% Tween-20 + 0.2% Triton X-100) and blocked for 1 hour with Protein Block (Spring Bioscience), incubated with primary antibodies overnight at 4°C, and washed with IHC buffer followed by incubation with respective secondary antibodies (Invitrogen) for 1 hour at room temperature.

Coverslip mounting was done using ProLong Diamond Gold Antifade Mountant with DAPI (Invitrogen) to stain nuclei. mages were acquired using Leica SP8 laser scanning confocal microscope. Quantification of the percentage of cells per mm^2^ labeled by NeuN or GFAP was based on three 10x magnification images per animal from a total of *n* = 4 mice. Cells within 100 µm of the ependymal layer (excluding the ependymal cells) in which DAPI signal was also evident were considered. All measurements were done using ImageJ.

For zebrafish, 6 dpf larvae were first blocked in blocking buffer (10 % Goat Serum, 1% BSA, 1% DMSO and 0.5 % Triton X-100 in PBS) overnight at 4°C, followed by incubation with primary antibodies, including mouse IgG2b anti-acetylated tubulin (1:200; Sigma-Aldrich) and anti-SV2 (1:200; DSHB), for 5 days with gentle agitation in a cold room. After washing 3 times with PBSTx, larvae were incubated with secondary antibodies, including goat anti-rabbit IgG Alexa Fluor 488 (1:500, Jackson ImmunoResearch) and goat anti-mouse IgG Alexa Fluor 647 antibody (1:500, Jackson ImmunoResearch, PA) for 3 days at 4°C. Following a series of washes, larvae were laterally mounted in 1.5% low melting point agarose (Sigma-Aldrich), and images were obtained using the Zeiss LSM-710 Confocal microscope.

### 7 T small animal MRI

Ten WT (3 males, 7 females, mean age = 114 days, *SD* = 12.9) and 10 KI (3 males, 7 females, mean age = 108 days, *SD* = 10.3) for a total of 20 mice were employed for high resolution, pre-clinical MR imaging experiments. Data from four animals were excluded from analysis to restrict subjects to an age range of approximately 3-4 months for consistency. For *in vivo* structural MRI, mice were anesthetized with isoflurane, placed in a plastic bed and restrained with gauze pads to minimize the possible influence of motion artifacts. For the duration of each MRI scan, mice were maintained under isoflurane gas anesthesia at approximately 37°C using a warm air blower and respiration was monitored using a pressure pad.

Imaging was performed using the 7 T Bruker Pharmascan (Bruker Biosciences, Billerica, MA), ultra-high field, pre-clinical MRI system of the McConnell Brain Imaging Centre at McGill University. The Pharmascan is equipped with an AVANCE II-model spectrometer and BFG-150/90-S shielded gradient system (Resonance Research Inc., Billerica, Massachusetts). Structural MR images were acquired using a 2D Rapid Imaging with Refocused Echoes (RARE) pulse sequence with the following parameters: effective echo time (TE_eff_): 30 ms, RARE factor: 8. In-plane resolution: 100 µm x 100 µm, slice thickness: 300 µm and receiver bandwidth: 46875 Hz. The repetition time (TR) and the number of acquired slices were varied for two pairs of WT/KI mice in order to achieve greater slice coverage along the rostro-caudal axis (TR: 4000 ms to 4750 ms, number of slices: 40 to 50). The number of averages was varied to optimize total scan time for mouse imaging under gas anesthesia. Lateral ventricles were manually segmented in each scan by a researcher blind to animal genotypes using the ITK-SNAP software (www.itksnap.org).^15^ and pooled lateral ventricle volumes were used for statistical analysis.

### Statistical Analysis

Continuous data were analyzed for normality and homogeneity of variance using Shapiro-Wilk tests (*n* < 50) or Kolmogorov-Smirnov (*n* ≥ 50) tests and Levene tests. Student’s *t*-tests or one-way ANOVAs were conducted when all assumptions were met. Welch’s t-tests were conducted when homogeneity of variance assumptions were not met. The nonparametric equivalent (Mann-Whitney U test) was conducted when normality assumptions were not met, or when both normality of data and homogeneity of variance assumptions were not met. A *p* value of < 0.05 was considered statistically significant. Data were analyzed using SPSS, R version 4.1.2 with Companion to Applied Regression package version 3.0, and Tidyverse version 1.3.1 software. All statistical analyses included multiple replicates from several independent experiments.

## Data availability

Raw data and statistical analyses for all major experiments are provided in Source Data. All identified spectra from the mass spectrometry experiment are available in Source Data, and raw proteomic data is available on PRIDE (PXD048699). Data concerning *in vitro* experiments and *in vivo* and *ex vivo* mouse experiments were generated at the Montreal Neurological Institute of McGill University. Data concerning zebrafish experiments were generated at the Oklahoma Medical Research Foundation. Derived data supporting any other findings of this study are available upon reasonable request from the corresponding author.

