## Supplementary Information for "Loss of symmetric cell division of apical neural progenitors drives *DENND5A*-related developmental and epileptic encephalopathy"

[†] The full list of authors in network is in Supplementary Notes.

#### Table of Contents

**Page**

1. **Supplementary Methods**
    a. **Motor skills scoring system……………………………………………….……..5**
    b. **Neurological phenotype scoring system…………………………….……..6**
    c. **Communication skills scoring system……………………………….……7**
    d. **Comorbidities scoring system………………………………………………...8**
2. **Supplementary Notes……………………………………………………………9-10**

### 1. Supplementary Methods

#### 1a. Motor skills scoring system

| Item | Scoring |
|---|---|
| Able to reach/grasp objects | **+1** if positive |
| Able to roll over | **+1** if positive |
| Able to sit with support | **+1** if positive OR is able to sit without support |
| Able to sit without support | **+1** if positive |
| Able to stand with support | **+1** if positive OR is able to stand without support |
| Able to stand without support | **+1** if positive |
| Able to walk with support | **+1** if positive OR is able to walk without support |
| Able to walk without support | **+1** if positive |
| Muscle tone or spasm problems | **+1** if negative for all (hyperreflexia, spastic tetraplegia, clonus, and current hyper/hypotonia) |
| Motor regression after seizure | **+1** if negative AND could perform one of the above behaviors in past |
| **TOTAL** | **10** |

Scoring system used for quantifying motor abilities. A low score reflects minimal motor abilities, a high score indicates a high degree of motor capabilities. If a child's ability to do a skill is unknown, it is counted as positive.

**1b. Neurological phenotype scoring system**

| **Item** | **Scoring** |
|---|---|
| Seizures | **+1** if positive |
| Reduced volume (cerebral or supratentorial parenchymal volume loss) | **+1** if positive |
| Cerebellum abnormalities (hypoplastic vermis, reduced volume) | **+1** if positive |
| Thalamus abnormalities (thalami fusion or reduced volume, massa intermedia prominence) | **+1** if positive |
| Basal ganglia abnormalities (dysplasia or reduced volume) | **+1** if positive |
| Calcifications | **+1** if positive |
| Ventricle or CSF abnormalities | **+1** if positive |
| White matter abnormalities (reduced corpus callosum or other white matter tract volume, delayed myelination or hyperintensity) | **+1** if positive |
| Hemorrhage or ischemic event | **+1** if positive |
| Cortical visual impairment | **+1** if positive |
| **TOTAL** | **10** |

Scoring system used for quantifying neurological phenotypes. A low score corresponds to few neurological abnormalities, a high score indicates many neurological abnormalities.

**1c. Communication skills scoring system**

| **Item** | **Scoring** |
|---|---|
| Smiles | **+1** if positive |
| Eye contact | **+1** if positive |
| Points at objects/people | **+1** if positive |
| Babbles | **+1** if positive OR if speaks in at least single words |
| Uses PECS board | **+1** if positive OR if speaks in at least single words |
| Speaks in single words | **+1** if positive OR if speaks in at least short phrases |
| Speaks in short phrases | **+1** if positive OR if speaks in sentences |
| Speaks in sentences | **+1** if positive |
| Language regression after seizure | **+1** if negative AND if had language skills in past |
| Receptive language delay | **+1** if negative AND at least babbles |
| **TOTAL** | **10** |

Scoring system used for quantifying communication abilities. A low score reflects minimal communication ability, a high score reflects more advanced language and communication abilities.

**1d. Comorbidities scoring system**

| Item | Scoring |
|---|---|
| Chronic constipation | **+1** if positive |
| Autism spectrum disorder (formally diagnosed or clinically suspected) | **+1** if positive |
| Psychiatric disorders (ADHD, anxiety) | **+1** if positive |
| Behavioral disorders or abnormalities (self-injury, poor sleep, hyperphagia) | **+1** if positive |
| Lung or breathing abnormalities (restrictive lung disease, asthma) | **+1** if positive |
| Cardiac abnormalities (ventricular/atrial septal defects, arrythmia) | **+1** if positive |
| Blindness | **+1** if positive |
| Obesity | **+1** if positive |
| Bone abnormalities (low density or osteoporosis, scoliosis, vertebral fusion, posterior fossa abnormality) | **+1** if positive |
| GERD | **+1** if positive |
| **TOTAL** | **10** |

Scoring system used for quantifying neurological phenotypes. A low score corresponds to few comorbidities, a high score indicates many comorbidities.

#### 2. Supplementary Notes

Members of the Undiagnosed Diseases Network include:

Maria T. Acosta, Margaret Adam, David R. Adams, Pankaj B. Agrawal, Mercedes E. Alejandro, Justin Alvey, Laura Amendola, Ashley Andrews, Euan A. Ashley, Mahshid S. Azamian, Carlos A. Bacino, Guney Bademci, Eva Baker, Ashok Balasubramanyam, Dustin Baldridge, Jim Bale, Michael Bamshad, Deborah Barbouth, Gabriel F. Batzli, Pinar Bayrak-Toydemir, Anita Beck, Alan H. Beggs, Edward Behrens, Gill Bejerano, Jimmy Bennet, Beverly Berg-Rood, Raphael Bernier, Jonathan A. Bernstein, Gerard T. Berry, Anna Bican, Stephanie Bivona, Elizabeth Blue, John Bohnsack, Carsten Bonnenmann, Devon Bonner, Lorenzo Botto, Brenna Boyd, Lauren C. Briere, Elly Brokamp, Gabrielle Brown, Elizabeth A. Burke, Lindsay C. Burrage, Manish J. Butte, Peter Byers, William E. Byrd, John Carey, Olveen Carrasquillo, Ta Chen Peter Chang, Sirisak Chanprasert, Hsiao-Tuan Chao, Gary D. Clark, Terra R. Coakley, Laurel A. Cobban, Joy D. Cogan, F. Sessions Cole, Heather A. Colley, Cynthia M. Cooper, Heidi Cope, William J. Craigen, Andrew B. Crouse, Michael Cunningham, Precilla D'Souza, Hongzheng Dai, Surendra Dasari, Mariska Davids, Jyoti G. Dayal, Matthew Deardorff, Esteban C. Dell'Angelica, Shweta U. Dhar, Katrina Dipple, Daniel Doherty, Naghmeh Dorrani, Emilie D. Douine, David D. Draper, Laura Duncan, Dawn Earl, David J. Eckstein, Lisa T. Emrick, Christine M. Eng, Cecilia Esteves, Tyra Estwick, Marni Falk, Liliana Fernandez, Carlos Ferreira, Elizabeth L. Fieg, Paul G. Fisher, Brent L. Fogel, Irman Forghani, Laure Fresard, William A. Gahl, Ian Glass, Rena A. Godfrey, Katie Golden-Grant, Alica M. Goldman, David B. Goldstein, Alana Grajewski, Catherine A. Groden, Andrea L. Gropman, Irma Gutierrez, Sihoun Hahn, Rizwan Hamid, Neil A. Hanchard, Kelly Hassey, Nichole Hayes, Frances High, Anne Hing, Fuki M. Hisama, Ingrid A. Holm, Jason Hom, Martha Horike-Pyne, Alden Huang, Yong Huang, Rosario Isasi, Fariha Jamal, Gail P. Jarvik, Jeffrey Jarvik, Suman Jayadev, Jean M. Johnston, Lefkothea Karaviti, Emily G. Kelley, Jennifer Kennedy, Dana Kiley, Isaac S. Kohane, Jennefer N. Kohler, Deborah Krakow, Donna M. Krasnewich, Elijah Kravets, Susan Korrick, Mary Koziura, Joel B. Krier, Seema R. Lalani, Byron Lam, Christina Lam, Brendan C. Lanpher, Ian R. Lanza, C. Christopher Lau, Kimberly LeBlanc, Brendan H. Lee, Hane Lee, Roy Levitt, Richard A. Lewis, Sharyn A. Lincoln, Pengfei Liu, Xue Zhong Liu, Nicola Longo, Sandra K. Loo, Joseph Loscalzo, Richard L. Maas, Ellen F. Macnamara, Calum A. MacRae, Valerie V. Maduro, Marta M. Majcherska, May Christine V. Malicdan, Laura A. Mamounas, Teri A. Manolio, Rong Mao, Kenneth Maravilla, Thomas C. Markello, Ronit Marom, Gabor Marth, Beth A. Martin, Martin G. Martin, Julian A. Martínez-Agosto, Shruti Marwaha, Jacob McCauley, Allyn McConkie-Rosell, Colleen E. McCormack, Alexa T. McCray, Elisabeth McGee, Heather Mefford, J. Lawrence Merritt, Matthew Might, Ghayda Mirzaa, Eva Morava-Kozicz, Paolo M. Moretti, Marie Morimoto, John J. Mulvihill, David R. Murdock, Mariko Nakano-Okuno, Avi Nath, Stan F. Nelson, John H. Newman, Sarah K. Nicholas, Deborah Nickerson, Donna Novacic, Devin Oglesbee, James P. Orengo, Laura Pace, Stephen Pak, J. Carl Pallais, Christina GS. Palmer, Jeanette C. Papp, Neil H. Parker, John A. Phillips III, Jennifer E. Posey, Lorraine Potocki, Barbara N. Pusey, Aaron Quinlan, Wendy Raskind, Archana N. Raja, Genecee Renteria, Chloe M. Reuter, Lynette Rives, Amy K. Robertson, Lance H. Rodan, Jill A. Rosenfeld, Natalie Rosenwasser, Robb K. Rowley, Maura Ruzhnikov, Ralph Sacco, Jacinda B. Sampson, Susan L. Samson, Mario Saporta, C. Ron

Scott, Judy Schaechter, Timothy Schedl, Kelly Schoch, Daryl A. Scott, Prashant Sharma, Vandana Shashi, Jimann Shin, Rebecca Signer, Catherine H. Sillari, Edwin K. Silverman, Janet S. Sinsheimer, Kathy Sisco, Edward C. Smith, Kevin S. Smith, Emily Solem, Lilianna Solnica-Krezel, Rebecca C. Spillmann, Joan M. Stoler, Nicholas Stong, Jennifer A. Sullivan, Kathleen Sullivan, Angela Sun, Shirley Sutton, David A. Sweetser, Virginia Sybert, Holly K. Tabor, Cecelia P. Tamburro, Queenie K.-G. Tan, Mustafa Tekin, Fred Telischi, Willa Thorson, Cynthia J. Tifft, Camilo Toro, Alyssa A. Tran, Brianna M.
